# Multivariate genome-wide association study (GWAS) of PTSD, Alcohol Use and Alcohol Use Disorders

**DOI:** 10.1101/2025.03.24.25324454

**Authors:** Peter B. Barr, Kaitlin Bountress, Chris Chatzinakos, James E. Hart, Zoe E. Neale, Christina Sheerin, Emma Johnson, Elizabeth G. Atkinson, Caroline M. Nievergelt, Adam X. Maihofer, Abigail Powers, Arpana Agrawal, Howard J. Edenberg, Joel Gelernter, Karestan C. Koenen, Bernice Porjesz, the PTSD workgroup of the Psychiatric Genomic Consortium (PGC-PTSD), the SUD workgroup of the Psychiatric Genomic Consortium (PGC-SUD), Ananda B. Amstadter, Jacquelyn L. Meyers

## Abstract

Alcohol use disorder (AUD) commonly co-occurs with posttraumatic stress disorder (PTSD), and comorbid PTSD and AUD is associated with poorer outcomes including worse treatment outcomes and significant physical health consequences. Both PTSD and AUD are polygenic in nature and genetically overlap. Previous work showed negative or non-significant associations between PTSD and alcohol consumption but positive genetic associations between PTSD and AUD. This work highlights the need for more nuanced examination of the similarities and distinctions in the associations between PTSD and alcohol consumption *versus* AUD. We leveraged the latest large-scale GWAS data to perform a multivariate GWAS of alcohol consumption (ALCC), problematic alcohol use (ALCP), and PTSD using GenomicSEM. Partial genetic correlations revealed that ALCP and PTSD were associated with each other (rG=0.39, *p =* 4.38×10^-60^) and with other psychiatric problems, medical conditions, and pain, while ALCC was generally only weakly correlated with PTSD (rG=-0.08, *p =* 2.03×10^-4^) and uncorrelated with most traits after accounting for its genetic overlap with ALCP and PTSD. We examined associations between GenomicSEM-derived polygenic scores (PGS) and their corresponding phenotypes in participants from the Collaborative Study on the Genetics of Alcoholism (COGA). PGS for ALCC were unrelated to PTSD diagnosis and PGS for PTSD were unrelated to drinks in a typical week. PTSD is more strongly related to alcohol problems, and much of the overlap in PTSD and consumption is accounted for by its overlap with alcohol problems. These results help demonstrate the complex partial overlaps of PTSD, AUD, and alcohol consumption.

## Introduction

Posttraumatic stress disorder (PTSD) is a debilitating condition occurring in an estimated 5.6% of individuals who have been exposed to trauma (1). Alcohol use disorder (AUD) commonly co-occurs with PTSD; a large epidemiological study found that 41.8% of those with PTSD also meet criteria for AUD (2). Comorbid PTSD and AUD is associated with poorer outcomes including higher symptom severity, decreased quality of life, poorer treatment outcomes, and significant physical health consequences (3). The increased risk for both elevated alcohol consumption and the development of AUD in individuals with PTSD aligns with the self-medication hypothesis that individuals with PTSD may use alcohol and other substances to dull the symptoms of PTSD (4). Conversely, the same study found that individuals who engage in risky drinking behaviors are more likely to be exposed to trauma (4). Another potential contribution to PTSD-AUD comorbidity is explored via the shared liability model, which posits that a common factor such as genetics may increase risk for both AUD and PTSD (5). Both PTSD and AUD are polygenic in nature, with results from twin studies estimating the heritability (*h*^2^) of PTSD at ∼30-40% (6–9) and AUD at ∼50% (10). Estimates of heritability from measured genotypic data, which estimate the amount of phenotypic variation explained by common single nucleotide polymorphisms (SNPs), are lower for each disorder, explaining 5-9% and 6-12% of the variance in PTSD and AUD (or their symptom scores), respectively (11–13).

Twin and family studies have demonstrated a latent genetic overlap between PTSD and AUD (14,15). These findings have largely been replicated using molecular genetic methods (i.e., linkage disequilibrium score regression, LDSC (16)) further supporting evidence of shared genetic overlap (17,18). Work by Bountress et al., 2022 (18) has also extended the investigation to include alcohol consumption, with findings indicating negative or non-significant associations between PTSD and alcohol use phenotypes (18), which differs from the positive associations between PTSD and AUD. Additionally, estimates of genetic correlations between problematic alcohol use and alcohol consumption phenotypes vary in strength, suggesting that these phenotypes are correlated but distinct (19,20). Together, these findings highlight the need for more nuanced examination of the overlaps among PTSD, AUD, and alcohol consumption.

Using multivariate genomic approaches (21), our group previously sought to enhance understanding of the relationship and pleiotropy among these phenotypes in samples most genetically similar to European reference panels (EUR-like) (18). Findings highlighted the importance of separating alcohol use from AUD. Specifically, removing genetic factors common to AUD and alcohol use resulted in an increase in each phenotype’s unique association with PTSD (i.e., PTSD was not correlated with the common factor, but was positively correlated with AUD, and was negatively correlated with alcohol use). These findings suggest that different genetic factors, likely those related to psychopathology, contribute to AUD uniquely compared to those important for alcohol use. Genetic factors related to alcohol use may be more genetically related to positive factors, such as life satisfaction (22). The landscape of psychiatric genetics is changing rapidly, and newer and more powerful GWAS summary statistics are increasingly available (23). Ongoing examination of these nuanced relationships, with the newer and larger samples, will provide greater clarity and support downstream gene discovery efforts.

In the current analyses we performed a multivariate genome wide association study (GWAS) using Genomic Structural Equation Modeling (GenomicSEM) (21). First, we replicated the structure of genetic overlap between PTSD, AUD, and alcohol consumption from prior GenomicSEM analyses (18), with increased statistical power from additional samples (11,12,19,24–26). Second, we examined the genetic correlations between these latent factors and relevant medical and behavioral traits to better understand the nature of shared and distinct genetic liability. Third, we applied a variety of post-GWAS techniques with available functional data to elucidate biological mechanisms, pathways, and tissues underlying these findings. Finally, we replicated genetic findings based on polygenic scores (PGS) derived from the latent PTSD, AUD, and alcohol consumption summary statistics in participants from the Collaborative Study on the Genetics of Alcoholism (COGA), an ancestrally diverse family sample enriched for alcohol use, misuse, and traumatic stress.

## Method

### Genomic Structural Equation Modeling (GenomicSEM)

We obtained summary statistics for PTSD and alcohol-related phenotypes using available large-scale GWAS described in Table 1. We limited our analyses to those classified as most genetically similar to European reference panels (EUR-like) due to the scarcity of large-scale summary statistics in other populations. GWAS for PTSD symptoms came from the PGC-PTSD Freeze 3, with COGA excluded (N = 1,307, 247) (12). PTSD case status reflects primarily lifetime PTSD diagnosis (i.e., controls have never had PTSD), but additionally current PTSD diagnosis (i.e., controls do not currently have PTSD) when lifetime diagnosis was unavailable. Indicators of our problematic alcohol use latent factor included GWASs of: a large scale meta-analysis of AUD, (N = 753,248) (11) with the COGA sample removed; the Alcohol Use Disorder Identification Test, or AUDIT (27), problems subscale from the UK Biobank (AUDIT-P, N = 160,824) (19); and maximum habitual alcohol use in a typical month from the Million Veteran Program cohort (MaxAlc, N = 218,623) (24). For alcohol consumption, we included GWAS of: drinks per week (DPW), defined as the average number of drinks a participant reported drinking each week, aggregated across all types of alcohol (N = 666,978; 23andMe excluded) (25); the AUDIT consumption subscale (AUDIT-C) in UK Biobank (N = 160,824) (19); and the AUDIT-C from MVP (N = 206,254) (26). The statistics used in the present analyses have gone through quality control pipelines applied by the specific consortia and are specified in the above corresponding publications. The analytic pipeline for the current analyses incorporated additional filtering. We standardized, cleaned, and aligned GWASs to the Genome Reference Consortium Human Build 37 through a common pipeline using the bcftools extension (28,29) of MungeSumstats (30).

**Table 1.** Descriptive Information about Included Phenotypes and Accompanying Cohorts.

| Phenotype | Accompanying GWAS | N | N effective | Primary Sample make-up | <u>Sex Distribution</u> |
| --- | --- | --- | --- | --- | --- |
| Alcohol Use Disorder | Zhou et al., 2023 | 753,248 | 352,373 | Civilian and Military | 67.9% Male |
| Max Habitual Alcohol Use | Deak et al., 2022 | 218,623 | 218,623 | Military | ~92.7% Male |
| AUDIT Problems | Mallard et al., 2022 | 160,824 | 160,824 | Civilian | ~43% Male |
| Drinks per Week | Saunders et al., 2022 | 666,978 | 666,978 | Civilian | NA |
| AUDIT Consumption | Mallard et al., 2022 | 160,824 | 160,824 | Civilian | ~43% Male |
| AUDIT Consumption | Kranzler et al., 2019 | 206,254 | 206,254 | Military | ~93% Male |
| PTSD Case/Control | Nievergelt et al., 2024 | 1,222,882 | 641,533 | Civilian and Military | ~52% Male |

Notes: PGC-SUD: Psychiatric Genomics Consortium Substance Use Disorder workgroup; MVP: Million Veteran Program, UKB: United Kingdom Biobank; GSCAN: GWAS & Sequencing Consortium of Alcohol and Nicotine use; PGC-PTSD: Psychiatric Genomics Consortium Posttraumatic Stress Disorder workgroup.

#### The Collaborative Study on the Genetics of Alcoholism (COGA)

COGA is a family-based, diverse sample of densely affected families for AUD and community controls (*N*_Individuals_ = 17,878; *N*_Family_ = 2,246; ∼25% self-identified African American, ∼52% female, ages 7-97 years (31)). All participants responded to the Semi-Structured Assessment of the Genetics of Alcoholism (SSAGA) (32) to assess: DSM-IV PTSD diagnosis (0: unaffected, 1: affected), frequency of alcohol use in a typical week (0: never, 1: once a week, 2: a few times a week, 3: nearly every day, 4: every day), and DSM-5 Alcohol Use Disorder diagnosis (0: unaffected, 1: affected). Genotyping was performed on 12,009 individuals using the Illumina 2.5M array, Illumina OmniExpress, Illumina 1M array, or the Affymetrix Smokescreen array; quality control and imputation have been previously described (33). Genetic similarity was assigned based on 1000 Genomes, phase 3 European and African reference panels. Participants were assigned to European (EUR-like, *N =* 8,038) and African (AFR-like, *N =* 3,654) groupings from genetic principal components (PCs) derived from GWAS data.

#### Model fitting and comparison

We conducted the multivariate GWAS using GenomicSEM, version 0.0.5c (21). First, we estimated genetic and sampling variance-covariance matrices using bivariate LDSC, which is robust to sample overlap (see *Supplemental Table 1* for genetic correlations among phenotypes). Second, we tested a series of structural equation models, attempting to minimize the discrepancy between the model-implied genetic covariance matrix and the empirical covariance matrix. We evaluated model fit using standard metrics, including the standardized root mean square residual (SRMR), model χ^2^, Akaike Information Criterion (AIC), and the Comparative Fit Index (CFI) (34,35). For case/control samples, we obtained liability scale estimates for SNP-*h*^2^ assuming a population prevalence of 10% for PTSD (36) and 16% for AUD. (37,38).

We estimated a series of four models and compared the fit of each (see Figure 1).

1. A common factor model (all 7 indicators [PTSD, AUD, AUDIT-P, MaxAlc, DPW, AUDIT-C [UKB], AUDIT-C[MVP]) loaded on a common factor, Figure 1A).
2. A two-factor model (alcohol-related GWAS loading on one factor and the PTSD GWAS loading on a second factor, Figure 1B).
3. A three-factor model with separate PTSD, alcohol consumption (ALCC), and alcohol problems (ALCP) factors (Figure 1C).
4. A bifactor model that allows for a factor common to all PTSD and alcohol phenotypes in addition to the phenotype specific factors described in the three-factor model. (Figure 1D).

**Figure 1:**
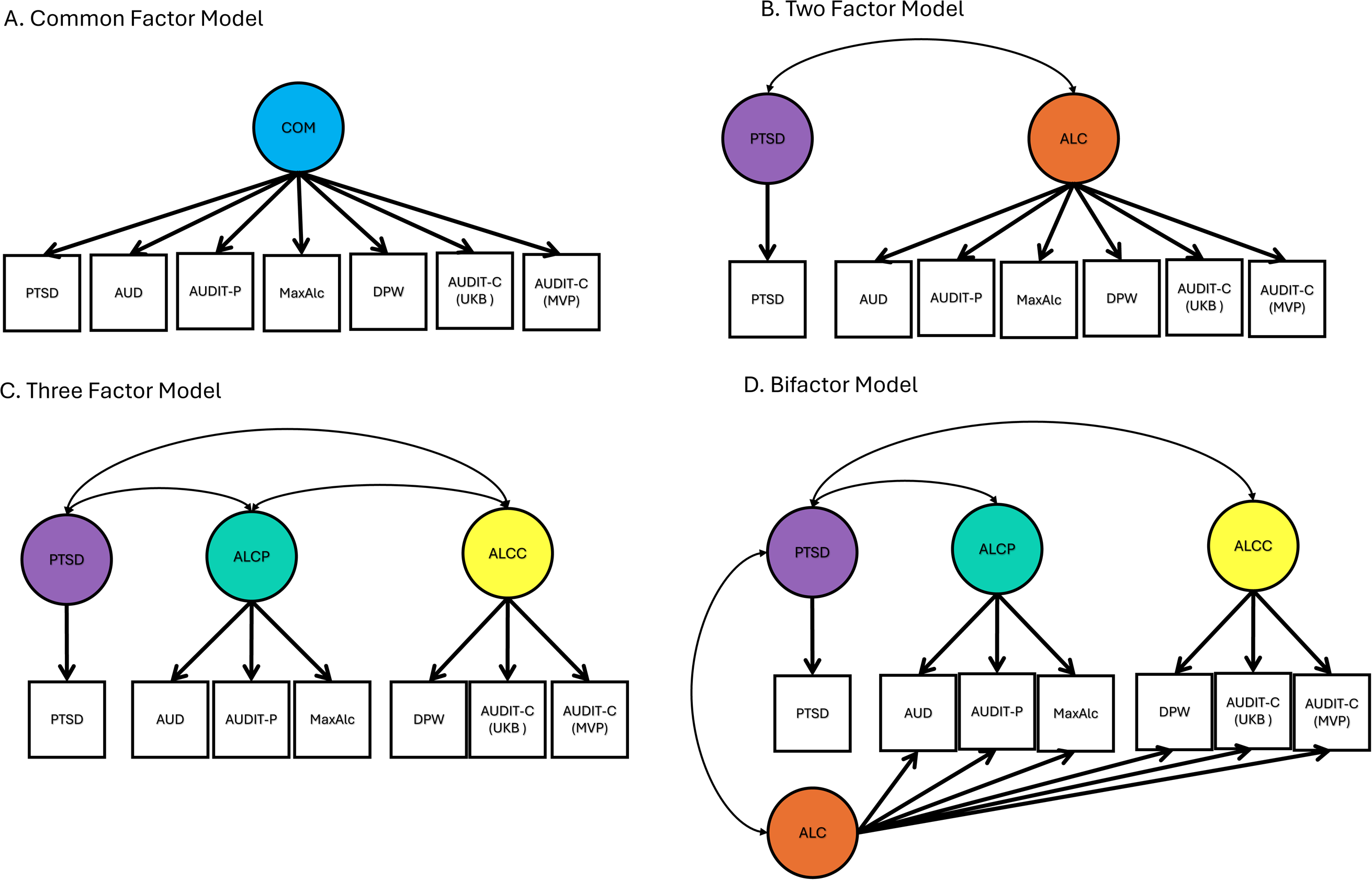
Tested Models of PTSD-ALC Relationship. Models tested for relationships between PTSD and alcohol-related outcomes. Models were fit in a stepwise manner beginning with A (common factor model) through D (bifactor model). We evaluated improvement in fit between models chi-square difference test.

To determine which model best fit the data, we examined the substantive interpretability of each model and its loadings, including the genetic associations between PTSD and factors common and specific to alcohol use and AUD. We also examined goodness-of-fit indices with the standard cut-offs for good fit, including a CFI: ≥ .9 and SRMR ≤ .08 and lower AIC values indicating better fit (39,40). We used the zero-order genetic correlations and prior work (18) to inform which alcohol items would load onto the ALCC-related factor (i.e., GSCAN drinks per week, UKB AUDIT-C, MVP AUDIT-C) or the ALCP-related factor (i.e., MVP AUDIT-Problems, MVP MaxAlc, Meta-Analysis of Problematic Alcohol Use). Ultimately in balancing model fit and interpretability, we chose the three factor model in analyses that follow.

#### Multivariate Genome Wide Association Study (GWAS)

After identifying the model with the combined greatest interpretability and best fit, we conducted a multivariate GWAS, calculating effect sizes for individual SNPs on each of the latent factors from that model. We also tested for heterogeneity in SNP effect sizes (QSNP) to investigate whether SNP associations were consistent with the implied factor structure, and gene-based tests of the QSNP statistics indicating whether heterogeneous effects clustered within genes. Additionally, we estimated genetic correlations between each of the three latent factors and multiple other indices of psychopathology, behavior and physical health outcomes, simultaneously. Evaluating ALCP, ALCC and PTSD genetics in tandem allows us to calculate the partial genetic correlation for each factor while accounting for the other two.

#### Post GWAS Analyses

We used multiple downstream *in silico* approaches (see the Supplementary Methods) to interpret the GWAS results of each of the latent factors: FUMA (FUMA version 1.3.6, (41) to define the associated genomic loci and prioritize implicated genes; MAGMA (version 1.08, (42)) to aggregate the association statistics for individual SNPs into tests of enrichment within protein-coding genes, and JEPEGMIX2-P (43) for a Transcriptome-Wide Association Study (TWAS); pathway analysis across 13 GTEx brain tissues, and Gene set enrichment analysis (GSEA) (44,45) to test concordance of differential expression analysis results with gene sets. A description of these methods is presented in the Supplemental Information.

#### Polygenic replication

We constructed polygenic scores (PGS) in COGA using PRS-CSx, which applies a Bayesian regression with a continuous shrinkage parameter to GWAS summary statistics from multiple populations simultaneously (46,47). We lacked sufficient GWAS in AFR-like population for multivariate GWAS, instead we used univariate GWAS of PTSD, AUD, and alcohol consumption to match to the latent factor GWASs from GenomicSEM. PGS were computed separately for EUR-like and AFR-like participants. We examined the association between the latent-factor derived PGS simultaneously with three primary outcomes in the COGA sample using a linear mixed model: DSM-IV PTSD diagnosis, DSM-5 Alcohol Use Disorder diagnosis, and frequency of alcohol use in a typical week. We included relatedness (i.e., family ID as random intercept), age, sex, 10 genetic principal components, and genotype array as covariates. After running the stratified analyses, we meta-analyzed the results using a fixed-effects meta-analysis.

## Results

### Model fitting

Table 2 presents the comparisons in model fit (see Supplemental Figure 1 for correlations among indicators). Neither the common factor model (Figure 1A; i.e., all 7 indicators loading on a single factor) nor the two-factor model (Figure 1B; i.e., Alcohol combined and PTSD factors) fit the data well. These models fit identically due to the same number of parameters tested in each (i.e., the rG between the second alcohol factor and PTSD vs no residual for PTSD, as it is set to 0). The three-factor model (Figure 1C) fit adequately χ [12]=299.69, AIC=331.69, CFI=.93, SRMR=.09), improving over the two-factor model. There was a moderate positive, genetic correlation between PTSD and ALCP (rG = .39, p = 4.4×10^-60^). The negative association between PTSD and ALCC was significant but small (rG = −.08, p = 2.0x 10^-4^), and the positive association between ALCP and ALCC was large (rG = .75, p < 5×10^-300^). Lastly, we fit a bifactor model (Figure 1D). The correlations between the common alcohol factor and ALCC specific and ALCP specific factors were fixed to zero, but the correlations between the PTSD factor and each of the three alcohol factors were freely estimated. Model D also fit the data well (χ^2^[6]=68.59, AIC=112.59, CFI=.98, SRMR=.04). The PTSD factor was positively correlated with the ALCP factor (rG = .53, p = 3.6×10^-5^), negatively correlated with the ALCC factor (rG = −.55, p = 5.3×10^-14^), and uncorrelated with the common alcohol factor (rG = .14, p = 0.13). Given the computational burden of running the bifactor model at scale and the issues with interpretability in bifactor models (48,49), we carried the three-factor model (Model C) forward to be used in the multivariate GWAS. All model estimates are presented in Supplemental Table 1.

**Table 2.**
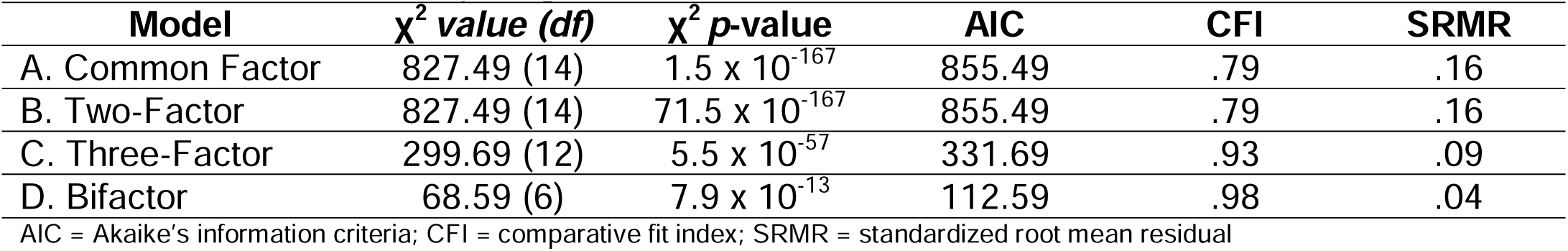
Fit indices for Competing GenomicSEM Models.

### Multivariate GWAS and post-GWAS analyses

Figure 2 presents the final model fit results and the Manhattan plots for each of the latent factors. After filtering for QC and SNPs in common across the seven indicators there were 3,799,881 SNPs (MAF > 1%) remaining in the multivariate GWAS. There were 52 loci associated with the PTSD factor, 94 loci associated with the ALCP factor, and 109 loci associated with the ALCC factor. Notably, there was very little overlap in individual loci across the three factors (Supplemental Figure 2), with no individual SNP being genome wide significant (*p* < 5×10^-8^) across all three factors, two SNPs shared across PTSD and ALCP (rs7519259, rs7629352), and four SNPs shared across ALCP and ALCC (rs1260326, rs13107325, rs2098112, rs6265, see Supplemental Table 3 for comparisons). Importantly, using a liberal threshold of .05/number of loci (a more conservative test of heterogeneity) for ALCC and ALCP, only 2 and 6 of the 109 and 94 loci, respectively, were significant in the Q-SNP analyses (full results in Supplemental Table 2), suggesting that many of the associations are best captured by the latent factor.

**Figure 2:**
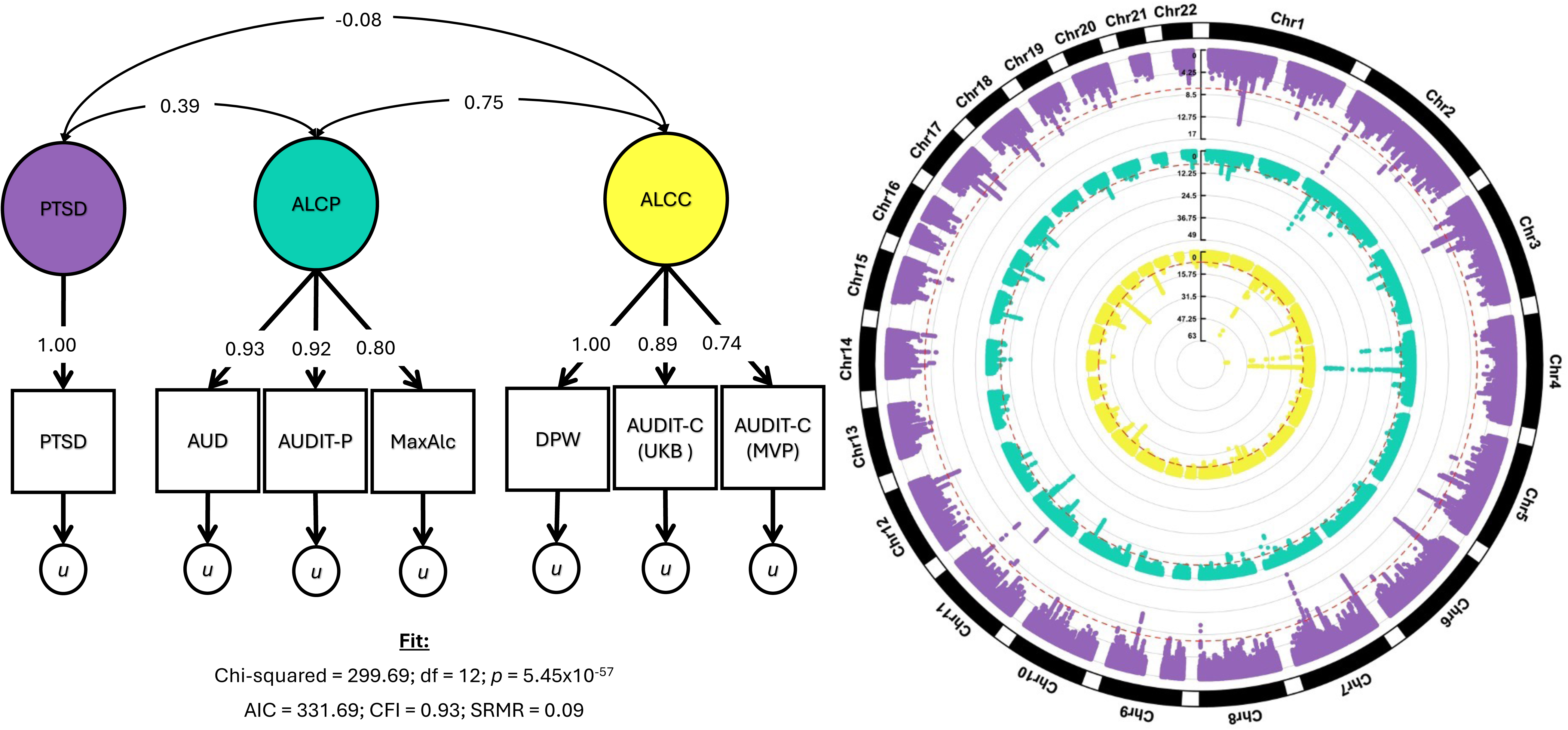
Model Fit and Multivariate Manhattan Plots. Panel A presents the final model used in the multivariate GWAS and corresponding factor loadings, factor correlations, and fit criteria. Panel B presents the circular Manhattan plots for the PTSD (purple), ALCP (teal), and ALCC (yellow) GenomicSEM results.

Gene-based analyses (via MAGMA) produced 132, 248, and 311 significant gene-based results for the PTSD, ALCP, and ALCC factors, respectively (Supplemental Tables 4 - 6). There was greater overlap among findings at the gene-based level; 14 genes were shared across all three factors, 23 genes across PTSD and ALCP, 103 genes across ALCP and ALCC, and 4 genes across PTSD and ALCC. While the majority of these 14 overlapping genes have previously been associated with either PTSD phenotypes, alcohol consumption or alcohol problems individually, 3 genes were novel: *NSF* (N-ethylmaleimide sensitive factor, vesicle fusing), *SPPL2C* (signal peptide peptidase like 2C), and *ARL17A* (ARF like GTPase 17A). The full gene-based results are presented in Supplemental Tables 4-6.

Figure 3 presents the partial genetic correlations across all three factors and related traits. As expected, the PTSD and ALCP factors were highly associated with commonly comorbid conditions (e.g., suicidal behaviors, depression, anxiety, opioid and cannabis use disorder), whereas the ALCC factor was not. The largest correlations for the PTSD factor were with PTSD sub-scales, suicidal behaviors and depression, whereas the largest correlations for the ALCP factor were with opioid and cannabis use disorder, as well as suicide attempts. For the ALCC factor, there were much lower or null correlations with many of the above mentioned psychiatric and substance use disorders, while there were substantial correlations with risk taking behaviors (which included alcohol use, full results in Supplemental Table. 7)

**Figure 3:**
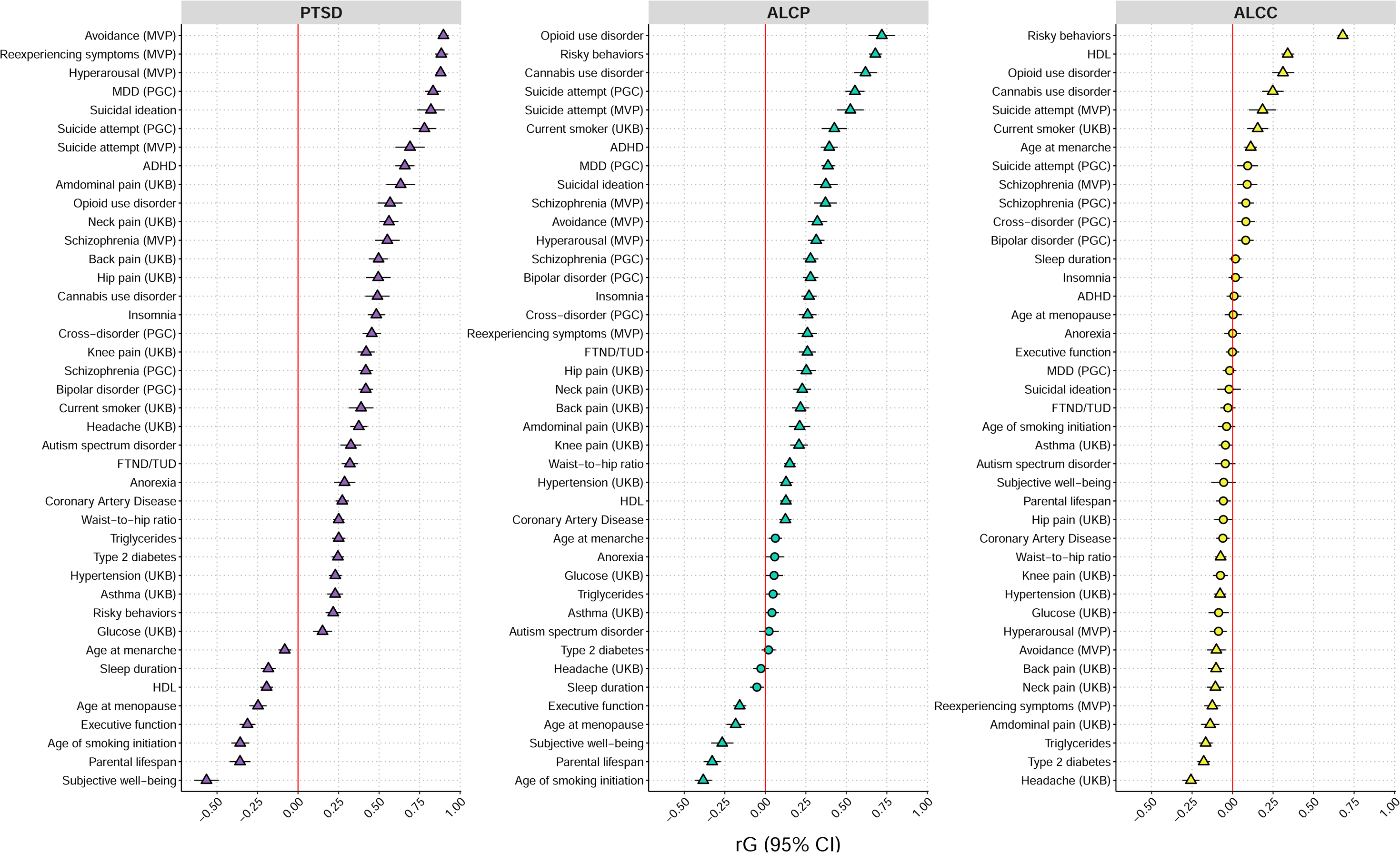
Genetic correlations between multivariate GWAS results and external phenotypes. Patial genetic correlations between each of the latent factor GWAS and selected external traits. Traits are ordered from strongest positive to strongest negative association within each facet. Triangles indicate genetic correlations are significant after adjusting for multiple testing, circles indicate not significant after correcting for multiple testing.

Next, we examined significant associations between GReX (genetically-regulated expression) in various brain regions and traits related to PTSD and alcohol use phenotypes via TWAS (Supplemental Table 8). Key regions, including the cortex, cerebellum, and subcortical structures (e.g., caudate, putamen), were implicated across all three latent factors (PTSD, ALCP, and ALCC) suggesting that these brain areas may play a central role in the shared genetic architecture underlying these conditions (Supplemental Figure 7). Further significant pathway analysis (FDR < 0.05) across 13 GTEx brain tissues for both the GenomicSEM derived PTSD factor and the original PTSD GWAS highlights key pathways enriched in multiple regions (Supplemental Figures 8 and 9). The frontal cortex, anterior cingulate cortex, and basal ganglia, including the putamen and nucleus accumbens, consistently show pathway intersections, suggesting these brain areas play a central role in the genetic architecture of PTSD. Additionally, the comparison of pathways across 13 GTEx brain tissues that overlap between the PTSD factor with the original PTSD GWAS reveal both shared and unique genetic mechanisms (Supplemental Figures 10). Key brain regions, including the anterior cingulate cortex, caudate, and cerebellum, demonstrate substantial overlap in pathways, suggesting common genetic influences. However, there are notable differences: the original PTSD GWAS shows more condition-specific pathways in regions like the caudate and cortex, while the GenomicSEM derived PTSD GWAS has unique pathways in the putamen and cerebellum. Finally, the Fisher exact test identifies significant pathways (FDR < 0.05) for PTSD-SUD across nine functional categories, ranging from Hallmark Gene Sets that reveal core biological processes to Cell Type Signature Gene Sets focused on cell-specific gene expressions (Supplemental Figure 11). Key brain regions, such as the anterior cingulate cortex, caudate, and putamen, are prominent across multiple categories, indicating their significant role in the genetic architecture of PTSD and alcohol use behavior. Additionally, categories like Oncogenic Signature Gene Sets and Immune-Related Gene Sets are notable, suggesting pathways related to cancer biology and immune system function may contribute to shared PTSD and alcohol use behavior risk. This comprehensive categorization offers valuable insights into the diverse biological mechanisms underlying these conditions.

### Polygenic Replication in COGA

Finally, we tested polygenic scores derived from the three latent factors for association with 1) number of drinks consumed in a typical week, 2) DSM-5 Alcohol Use Disorder, and 3) DSM-IV PTSD in COGA participants of European-like and African-like ancestry. Figure 4 presents the meta-analyzed results. The PTSD-factor PGS was associated with PTSD diagnosis (Beta = 0.057, SE = 0.012, *p =* 1.00×10^-6^) and AUD diagnosis (Beta = 0.036, SE = 0.011, *p =* 1.14×10^-3^), but not with typical drinks per week (Beta = 0.002, SE = 0.011, *p =* 8.25×10^-1^). Likewise, the ALCC-factor PGS was associated with typical drinks per week (Beta = 0.052, SE = 0.010, *p =* 4.63×10^-8^) and AUD diagnosis (Beta = 0.076, SE = 0.010, *p =* 3.98×10^-15^), but not PTSD (Beta = 0.021, SE = 0.010, *p =* 3.74×10^-2^). However, The ALCP-factor PGS was associated all three outcomes: AUD diagnosis (Beta = 0.118, SE = 0.010, *p* = 1.91×10^-35^), PTSD diagnosis (Beta = 0.036, SE = 0.010, *p* = 2.80×10^-4^), and typical drinks per week (Beta = 0.086, SE = 0.010, *p* = 3.65×10^-19^). The stratified and meta-analysis results are presented in Supplemental Table 9.

**Figure 4:**
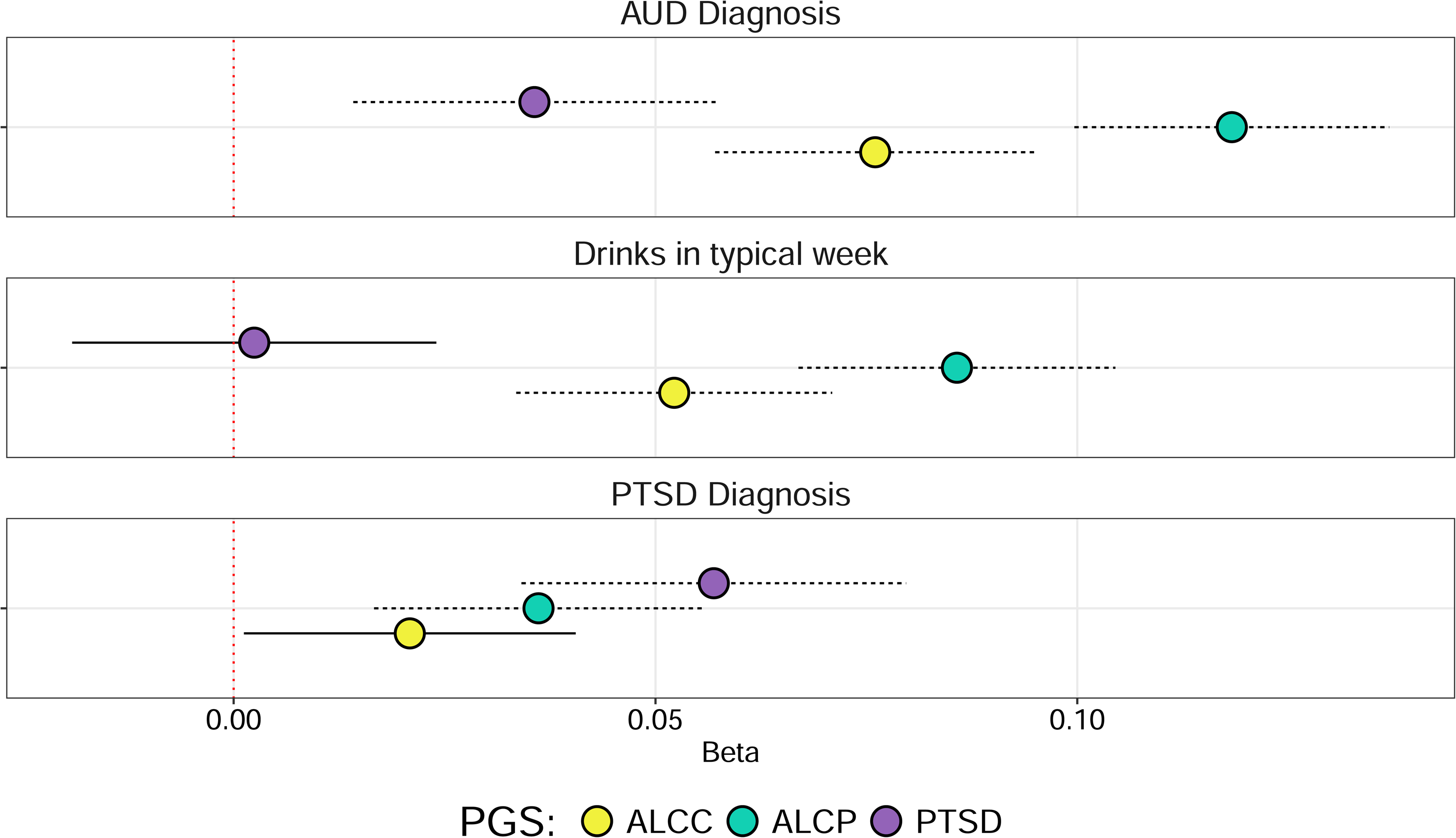
Polygenic score (PGS) associations in the Collaborative Study on the Genetics of Alcoholism (COGA) Conditional betas (and 95% confidence intervals) for associations between polygenic scores for PTSD (purple), ALCP (teal), ALCC (yellow) and corresponding outcomes in COGA. Each PGS was included simultaneously in the model. Dotted lines indicate significant associations after correcting for multiple testing.

## Discussion

This study leveraged existing large-scale GWAS data to examine the genetic architecture of alcohol consumption, AUD, and PTSD using GenomicSEM to expand genetic discovery through multivariate GWAS (21). This approach produced both novel results and replicated previously identified loci associated with each of these phenotypes. We further elucidated genetic correlations between these GenomicSEM derived factors and other medically and behaviorally relevant traits, and applied available functional data to elucidate biological mechanisms, pathways, and tissues underlying these findings, highlighting 17q21.3 as a locus for shared genetic association across PTSD, alcohol use (i.e., drinks per week and AUDIT consumption), and alcohol problems (i.e., problematic alcohol use, AUDIT problems, and max alcohol use). While this locus is in a known inversion region and any specific mapped genes should be interpreted with caution; genes in this region have been implicated in prior transcriptome profiling of postmortem brain tissue of individuals with PTSD (50). We ran a series of robustness checks examining FDR significant genes by cytoband for each GWAS (Supplemental Figures 12-14) and 17q21.31 consistently emerged as a top result across each latent factor. Finally, we found that polygenic scores based on these GenomicSEM derived PTSD, ALCP, and ALCC genetic factors replicated in participants from the Collaborative Study on the Genetics of Alcoholism (COGA).

Our GenomicSEM models, using the most up to date and well-powered GWAS summary statistics build upon prior work (18). Consistent with prior findings, our results suggest that there is a qualitative difference between alcohol use phenotypes (e.g., drinks per week) and more severe alcohol problems (e.g., AUD) regarding their genetic relationships with PTSD, with this prior work finding that the genetic effects on these phenotypes are distinct (18,19,51,52). Specifically, the ALCP factor was moderately positively correlated with PTSD, whereas the ALCC factor was weakly negatively correlated with PTSD. The consistency of the results of the GenomicSEM models between prior work and the present analysis is reassuring, and underscores the need to examine the alcohol consumption separately from that of problematic alcohol use. As sample sizes have expanded rapidly, gene identification has increased for PTSD and alcohol phenotypes individually (11,12). Here we built on this prior work to consider how the genetic influence on these phenotypes may be shared, mimicking other recent multivariate GWAS demonstrating the utility of this approach (53).

The multivariate GWAS identified 52 genomic risk loci for the PTSD factor, 94 genomic risk loci for the Alcohol Problems factor, and 109 genomic risk loci for the alcohol consumption factor. While there was little overlap in individual loci across the three factors, which is not inconsistent with prior work (51), there were two SNPs shared across PTSD and alcohol problems (rs7519259, rs7629352), and four SNPs shared across alcohol problems and alcohol consumption (rs1260326, rs13107325, rs2098112, rs6265). It is notable that rs1229984 in *ADH1B*, the strongest association in alcohol GWAS to date, was associated with problems but not consumption in the multivariate GWAS. However, this is consistent with recent multivariate GWAS of alcohol phenotypes, which also found rs1229984 loaded exclusively on a problems factor and not with other consumption-related factors (54). Interestingly, rs7519259, an intronic variant in Phosphodiesterase 4B (*PDE4B*) located on 1p31.3, has not previously been associated with either PTSD or alcohol use problems but *has* been associated with general addiction (i.e., capturing alcohol, cannabis, opioid use disorder) (55) and BMI (56). This gene is also a binding site for *DISC1* (disrupted in schizophrenia 1). Similarly, rs7629352, an intronic phospholipase C-like 2 (*PLCL2*) variant located on 3p24.3, associated with ALCP here, has previously been associated with smoking initiation (25) but not PTSD or alcohol use behavior. *GCKR* variant rs1260326, *SLC39A8* variant rs13107325, and *DPP6* variant rs2098112 have all been robustly associated with both alcohol consumption and alcohol use disorder (24,57,58). *BDNF* variant rs6265 has been associated with substance-related or substance-adjacent traits (e.g., smoking behavior (59), risk taking behavior (60), and BMI related traits (61)), but not explicitly with alcohol consumption or alcohol use disorder.

When comparing the number of signals identified via multivariate GWAS of the GenomicSEM-derived factors to the original GWAS of each related phenotype, we found that GenomicSEM-derived alcohol consumption and alcohol problems factors performed comparably or out-performed the original GWAS. For example, the original GWAS of alcohol problems produced 2-110 genome-wide significant loci, whereas the GenomicSEM factor produced 94 loci; the original GWAS of alcohol consumption produced 8-13 genome-significant loci, whereas the GenomicSEM factor produced 109 loci. This suggests that leveraging the gSEM derived factors improved gene/locus identification efforts. The original PTSD results in EUR-like populations out-performed the multivariate GWAS--76 loci in the original (81 loci, of which 5 were on X which was not included here) compared with 52 in the GenomicSEM derived factor. Of the 52 loci, 27 were the same as those in the original GWAS, while the remaining 25 were SNPs in high LD (*r^2^* > 0.6) with loci in the original PTSD GWAS. When we filtered the original PTSD results to the SNPs included in the GenomicSEM model (3,799,881 vs. 6,545,577) we identified 62 loci using the same LD threshold in FUMA, suggesting only part of the reduction in loci is explained by the reduction to SNPs in common for all indicator GWAS required when using GenomicSEM (see Supplemental Table 10 for the 62 loci from the original results available in the PTSD-factor). In total, although we identified fewer loci than the original PTSD GWAS, these associations can be viewed, in part, as those specific to PTSD, with the covariance of PTSD and the alcohol factors removed.

Gene-based analyses produced 132, 248, and 311 genome-wide significant gene-based results for the PTSD-factor, ALCP-factor, and ALCC-factor respectively. There was greater overlap among findings at the gene-based level; 14 genes were shared across all three factors, 23 genes across PTSD and ALCP, 103 genes across ALCP and ALCC, and 4 genes across PTSD and ALCC. While the majority of the 14 genes that were GWS for PTSD, ALCP and ALCC have previously been associated with either PTSD disorder, alcohol consumption or alcohol problems individually, there were 2 genes that were novel. *NSF* (N-ethylmaleimide sensitive factor, vesicle fusing) 17q21.3 and *SPPL2C* (signal peptide peptidase like 2C located on 17q21.3)—both of which have been previously associated with relevant traits (e.g., *NSF—* anxiety, worry, executive functioning, Alzheimer’s; *SPPL2C—*ADHD, Alzheimer’s, Parkinson’s). Additionally, *ARL17B,* which prior analyses have shown to be upregulated in post mortem brain tissue for those with PTSD (50), was also is upregulated in multiple cell types in carriers of a missense variant of *SPPL2C* in analyses of Alzheimer’s disease (62).

Partial genetic correlations between the 3 factors and medically-and behaviorally-relevant traits were generally as expected (63–65), with the PTSD and ALCP factors being highly associated with commonly comorbid conditions (e.g., depression, anxiety, suicidal behaviors, bipolar disorder, substance use disorders), whereas the alcohol consumption factor was not. For the ALCC factor, there were much lower or null correlations with the above mentioned psychiatric and substance use disorders, while there were substantial correlations with risk taking behaviors. Interestingly the genetic correlation with ALCC and educational attainment was in the opposite direction for the same correlation with either ALCP or PTSD, suggesting GWAS of alcohol consumption are still capturing some residual socioeconomic confounding. Other interesting findings pertained to the significant genetic associations between our phenotypes and different types of pain (e.g., neck, knee, hip, abdominal), with the associations being significant and positive for ALCP and PTSD, but significant and negative for ALCC. There very limited work finding significant positive genetic correlations between PTSD and pain (66), and some preliminary GWAS that suggest some hits may be shared by AUD and chronic pain (67). More work is needed to clarify the direction of genetic effects between these constructs, as well as whether there might be shared influences that directly impact on this comorbidity.

Additionally, we examined significant associations between gene expression in various brain regions and traits related to PTSD and AUD via TWAS. Key regions, including the cortex, cerebellum, and subcortical structures (e.g., caudate, putamen), were implicated across all three GenomicSEM factors (PTSD, ACLP, ALCC) suggesting that these brain areas may play a central role in the shared genetic architecture underlying these conditions. The comparison of pathways across 13 GTEx brain tissues that overlap between the GenomicSEM factors with the original GWAS reveal both shared and unique genetic mechanisms. Key brain regions, including the anterior cingulate cortex, caudate, and cerebellum, demonstrate substantial overlap in pathways, suggesting common genetic influences. However, there are notable differences: the original PTSD GWAS shows more condition-specific pathways in regions like the caudate and cortex, while the GenomicSEM derived PTSD GWAS has unique pathways in the putamen and cerebellum. This interplay between shared and unique genetic architectures across brain regions highlights the commonalities and distinctions in the molecular underpinnings of these PTSD “subtypes” (PTSD as a whole vs PTSD with shared variance with alcohol use behavior and alcohol problems removed). Key brain regions, such as the anterior cingulate cortex, caudate, and putamen, are prominent across multiple categories, indicating their significant role in the genetic architecture of PTSD-AUD. Additionally, categories like Oncogenic Signature Gene Sets and Immune-Related Gene Sets identified in the PTSD factor are notable, suggesting pathways related to cancer biology and immune system function may contribute to shared PTSD and alcohol use behavior risk. This comprehensive categorization offers valuable insights into the diverse biological mechanisms underlying these conditions.

We created polygenic scores derived from the three GenomicSEM factors in COGA participants of European-like and African-like genetic similarity. The COGA sample is unique in its ascertainment for those with AUD and their relatives, in addition to community-based comparison families (68). The rates of alcohol use and AUD in COGA are relatively high compared with the general population, as is exposure to traumatic stress. Each GenomicSEM based-PGS was associated with its component phenotype (i.e., the PTSD PGS was associated with PTSD, the alcohol problems PGS was associated with DSM-5 AUD, and the Alcohol Consumption PGS was associated with number of drinks consumed in a typical week. In addition, several cross-associations were observed (e.g., the PTSD PGS was associated with DSM-5 AUD, but not typical drinks per week). Findings point to the overlap in AUD and PTSD, but not PTSD and alcohol consumption, reiterating key results from the multivariate GWAS and corresponding partial genetics correlations.

There are several important limitations of the current research. The primary limitation is that discovery samples were derived from European-like participants alone. While we tried to increase representation in the polygenic score analyses, there is still a large amount of work to be done before GWAS reach parity across populations. Second, our PTSD factor only had one indicator and this approach comes with the assumption that our PTSD indicator perfectly captures all variance for PTSD. As new well-powered GWAS are conducted and made available, future work should expand the number of indicators to enable more precise estimations. Lastly, we explored the overlap of PTSD, alcohol problems, and alcohol consumption averaged across the life course. Expanding the current results into data that allows us to explore these phenomena prospectively may allow us to further tease apart the nature of the relationship. Lastly, while the bivariate model had superior fit statistics, the three-factor model was used for our multivariate analyses.

In conclusion, this study leveraged the largest existing large-scale GWAS data to capture the shared and unique genetic relationships among alcohol consumption, AUD, and PTSD using GenomicSEM, ultimately expanding novel gene discovery using multivariate GWAS. This produced both novel results and replicated previously identified loci associated with each of these phenotypes. We further elucidated biological mechanisms, pathways, and tissues underlying these findings highlighting 17q21.3 as a locus for shared genetic association across PTSD and alcohol use and alcohol problems, and replicated findings via polygenic scores in participants from the Collaborative Study on the Genetics of Alcoholism.

## Data Availability

Information on availability of GWAS summary statistics are listed in their corresponding p publications. COGA is available through dbGaP (accession number: phs000763.v1.p1)

## Acknowledgments

This study was supported in part by the National Institute of Drug Abuse (R01DA060596, R01DA054138), and the National Institute on Alcohol Abuse and Alcoholism (U10AA008401, R01AA030549), and NIMH R01MH106595. AA was additionally supported by R01DA054869. KB was supported by K01AA028058 and R34DA061267. The content of this article is solely the responsibility of the authors and does not necessarily represent the official views of the National Institutes of Health.

## References

1. Koenen KC, Ratanatharathorn A, Ng L, McLaughlin KA, Bromet EJ, Stein DJ, et al. Posttraumatic stress disorder in the World Mental Health Surveys. Psychol Med. 2017 Oct;47(13):2260–74.

2. Pietrzak RH, Goldstein RB, Southwick SM, Grant BF. Prevalence and Axis I comorbidity of full and partial posttraumatic stress disorder in the United States: results from Wave 2 of the National Epidemiologic Survey on Alcohol and Related Conditions. J Anxiety Disord. 2011 Apr;25(3):456–65.

3. Norman SB, Haller M, Hamblen JL, Southwick SM, Pietrzak RH. The burden of co-occurring alcohol use disorder and PTSD in U.S. Military veterans: Comorbidities, functioning, and suicidality. Psychol Addict Behav. 2018;32(2):224–9.

4. Hawn SE, Cusack SE, Amstadter AB. A Systematic Review of the Self-Medication Hypothesis in the Context of Posttraumatic Stress Disorder and Comorbid Problematic Alcohol Use. J Trauma Stress. 2020 Oct;33(5):699–708.

5. Danovitch I. Post-traumatic stress disorder and opioid use disorder: A narrative review of conceptual models. J Addict Dis. 2016 Jul 2;35(3):169–79.

6. Koenen KC, Hitsman B, Lyons MJ, Niaura R, McCaffery J, Goldberg J, et al. A twin registry study of the relationship between posttraumatic stress disorder and nicotine dependence in men. Arch Gen Psychiatry. 2005 Nov;62(11):1258–65.

7. Koenen KC, Lyons MJ, Goldberg J, Simpson J, Williams WM, Toomey R, et al. A high risk twin study of combat-related PTSD comorbidity. Twin Res Off J Int Soc Twin Stud. 2003 Jun;6(3):218–26.

8. True WR, Rice J, Eisen SA, Heath AC, Goldberg J, Lyons MJ, et al. A twin study of genetic and environmental contributions to liability for posttraumatic stress symptoms. Arch Gen Psychiatry. 1993 Apr;50(4):257–64.

9. Stein MB, Jang KL, Taylor S, Vernon PA, Livesley WJ. Genetic and environmental influences on trauma exposure and posttraumatic stress disorder symptoms: a twin study. Am J Psychiatry. 2002 Oct;159(10):1675–81.

10. Verhulst B, Neale MC, Kendler KS. The heritability of alcohol use disorders: a meta-analysis of twin and adoption studies. Psychol Med. 2015 Apr;45(5):1061–72.

11. Zhou H, Kember RL, Deak JD, Xu H, Toikumo S, Lind PA, et al. Multi-ancestry study of the genetics of problematic alcohol use in over 1 million individuals. Nat Med. 2023 Dec;29(12):3184–92.

12. Nievergelt CM, Maihofer AX, Atkinson EG, Chen CY, Choi KW, Coleman JRI, et al. Genome-wide association analyses identify 95 risk loci and provide insights into the neurobiology of post-traumatic stress disorder. Nat Genet. 2024 May;56(5):792–808.

13. Stein MB, Levey DF, Cheng Z, Wendt FR, Harrington K, Pathak GA, et al. Genome-wide association analyses of post-traumatic stress disorder and its symptom subdomains in the Million Veteran Program. Nat Genet. 2021 Feb;53(2):174–84.

14. Xian H, Chantarujikapong SI, Scherrer JF, Eisen SA, Lyons MJ, Goldberg J, et al. Genetic and environmental influences on posttraumatic stress disorder, alcohol and drug dependence in twin pairs. Drug Alcohol Depend. 2000 Dec 22;61(1):95–102.

15. McLeod DS, Koenen KC, Meyer JM, Lyons MJ, Eisen S, True W, et al. Genetic and Environmental Influences on the Relationship Among Combat Exposure, Posttraumatic Stress Disorder Symptoms, and Alcohol Use. J Trauma Stress. 2001 Apr 1;14(2):259–75.

16. Bulik-Sullivan BK, Loh PR, Finucane HK, Ripke S, Yang J, Patterson N, et al. LD Score regression distinguishes confounding from polygenicity in genome-wide association studies. Nat Genet. 2015 Mar;47(3):291–5.

17. Sheerin CM, Bountress KE, Meyers JL, de Viteri SSS, Shen H, Maihofer AX, et al. Shared molecular genetic risk of alcohol dependence and PTSD. Psychol Addict Behav J Soc Psychol Addict Behav. 2020 Aug;34(5):613–9.

18. Bountress KE, Brick LA, Sheerin C, Grotzinger A, Bustamante D, Hawn SE, et al. Alcohol use and alcohol use disorder differ in their genetic relationships with PTSD: A genomic structural equation modelling approach. Drug Alcohol Depend. 2022 May 1;234:109430.

19. Mallard TT, Savage JE, Johnson EC, Huang Y, Edwards AC, Hottenga JJ, et al. Item-Level Genome-Wide Association Study of the Alcohol Use Disorders Identification Test in Three Population-Based Cohorts. Am J Psychiatry. 2022 Jan;179(1):58–70.

20. Sanchez-Roige S, Fontanillas P, Elson SL, Team T 23andMe R, Gray JC, de Wit H, et al. Genome-wide association study of alcohol use disorder identification test (AUDIT) scores in 20 328 research participants of European ancestry. Addict Biol. 2019;24(1):121–31.

21. Grotzinger AD, Rhemtulla M, de Vlaming R, Ritchie SJ, Mallard TT, Hill WD, et al. Genomic structural equation modelling provides insights into the multivariate genetic architecture of complex traits. Nat Hum Behav. 2019 May;3(5):513–25.

22. Bountress KE, Cusack SE, Hawn SE, Grotzinger A, Bustamante D, Kirkpatrick RM, et al. Genetic associations between alcohol phenotypes and life satisfaction: a genomic structural equation modelling approach. Sci Rep. 2023 Aug 18;13(1):13443.

23. Abdellaoui A, Yengo L, Verweij KJH, Visscher PM. 15 years of GWAS discovery: Realizing the promise. Am J Hum Genet. 2023 Feb 2;110(2):179–94.

24. Deak JD, Levey DF, Wendt FR, Zhou H, Galimberti M, Kranzler HR, et al. Genome-Wide Investigation of Maximum Habitual Alcohol Intake in US Veterans in Relation to Alcohol Consumption Traits and Alcohol Use Disorder. JAMA Netw Open. 2022 Oct 27;5(10):e2238880.

25. Saunders GRB, Wang X, Chen F, Jang SK, Liu M, Wang C, et al. Genetic diversity fuels gene discovery for tobacco and alcohol use. Nature. 2022 Dec;612(7941):720–4.

26. Kranzler HR, Zhou H, Kember RL, Vickers Smith R, Justice AC, Damrauer S, et al. Genome-wide association study of alcohol consumption and use disorder in 274,424 individuals from multiple populations. Nat Commun. 2019 Apr 2;10(1):1499.

27. Saunders JB, Aasland OG, Babor TF, de la Fuente JR, Grant M. Development of the Alcohol Use Disorders Identification Test (AUDIT): WHO Collaborative Project on Early Detection of Persons with Harmful Alcohol Consumption--II. Addict Abingdon Engl. 1993 Jun;88(6):791–804.

28. Genovese G, Rockweiler NB, Gorman BR, Bigdeli TB, Pato MT, Pato CN, et al. BCFtools/liftover: an accurate and comprehensive tool to convert genetic variants across genome assemblies. Bioinforma Oxf Engl. 2024 Jan 2;40(2):btae038.

29. Genovese G. freeseek/score [Internet]. 2025. Available from: https://github.com/freeseek/score

30. Murphy AE, Schilder BM, Skene NG. MungeSumstats: a Bioconductor package for the standardization and quality control of many GWAS summary statistics. Bioinforma Oxf Engl. 2021 Dec 7;37(23):4593–6.

31. Johnson EC, Salvatore JE, Lai D, Merikangas AK, Nurnberger JI, Tischfield JA, et al. The collaborative study on the genetics of alcoholism: Genetics. Genes Brain Behav. 2023 Jun 30;22(5):e12856.

32. Bucholz KK, Cadoret R, Cloninger CR, Dinwiddie SH, Hesselbrock VM, Nurnberger JI, et al. A new, semi-structured psychiatric interview for use in genetic linkage studies: a report on the reliability of the SSAGA. J Stud Alcohol. 1994 Mar;55(2):149–58.

33. Lai D, Wetherill L, Bertelsen S, Carey CE, Kamarajan C, Kapoor M, et al. Genome-wide association studies of alcohol dependence, DSM-IV criterion count and individual criteria. Genes Brain Behav. 2019 Jul;18(6):e12579.

34. Browne MW. Asymptotically distribution-free methods for the analysis of covariance structures. Br J Math Stat Psychol. 1984 May;37 ( Pt 1):62–83.

35. Savalei V, Bentler PM. A Two-Stage Approach to Missing Data: Theory and Application to Auxiliary Variables. Struct Equ Model Multidiscip J. 2009 Jul 14;16(3):477–97.

36. Nievergelt CM, Maihofer AX, Klengel T, Atkinson EG, Chen CY, Choi KW, et al. International meta-analysis of PTSD genome-wide association studies identifies sex- and ancestry-specific genetic risk loci. Nat Commun. 2019 Oct 8;10(1):4558.

37. Duncan LE, Ratanatharathorn A, Aiello AE, Almli LM, Amstadter AB, Ashley-Koch AE, et al. Largest GWAS of PTSD (N=20LJ070) yields genetic overlap with schizophrenia and sex differences in heritability. Mol Psychiatry. 2018 Mar;23(3):666–73.

38. Walters RK, Polimanti R, Johnson EC, McClintick JN, Adams MJ, Adkins AE, et al. Transancestral GWAS of alcohol dependence reveals common genetic underpinnings with psychiatric disorders. Nat Neurosci. 2018 Dec;21(12):1656–69.

39. Hu L, Bentler PM. Cutoff criteria for fit indexes in covariance structure analysis: Conventional criteria versus new alternatives. Struct Equ Model Multidiscip J. 1999 Jan 1;6(1):1–55.

40. Kenny DA, Kaniskan B, McCoach DB. The performance of RMSEA in models with small degrees of freedom. Sociol Methods Res. 2015;44(3):486–507.

41. Watanabe K, Taskesen E, van Bochoven A, Posthuma D. Functional mapping and annotation of genetic associations with FUMA. Nat Commun. 2017 Nov 28;8(1):1826.

42. de Leeuw CA, Mooij JM, Heskes T, Posthuma D. MAGMA: generalized gene-set analysis of GWAS data. PLoS Comput Biol. 2015 Apr;11(4):e1004219.

43. Chatzinakos C, Georgiadis F, Lee D, Cai N, Vladimirov VI, Docherty A, et al. TWAS pathway method greatly enhances the number of leads for uncovering the molecular underpinnings of psychiatric disorders. Am J Med Genet Part B Neuropsychiatr Genet Off Publ Int Soc Psychiatr Genet. 2020 Dec;183(8):454–63.

44. Subramanian A, Tamayo P, Mootha VK, Mukherjee S, Ebert BL, Gillette MA, et al. Gene set enrichment analysis: A knowledge-based approach for interpreting genome-wide expression profiles. Proc Natl Acad Sci. 2005 Oct 25;102(43):15545–50.

45. Mootha VK, Lindgren CM, Eriksson KF, Subramanian A, Sihag S, Lehar J, et al. PGC-1α-responsive genes involved in oxidative phosphorylation are coordinately downregulated in human diabetes. Nat Genet. 2003 Jul;34(3):267–73.

46. Ge T, Chen CY, Ni Y, Feng YCA, Smoller JW. Polygenic prediction via Bayesian regression and continuous shrinkage priors. Nat Commun. 2019 Apr 16;10(1):1776.

47. Ruan Y, Lin YF, Feng YCA, Chen CY, Lam M, Guo Z, et al. Improving polygenic prediction in ancestrally diverse populations. Nat Genet. 2022 May;54(5):573–80.

48. Bonifay W, Lane SP, Reise SP. Three concerns with applying a bifactor model as a structure of psychopathology. Clin Psychol Sci. 2017;5(1):184–6.

49. Dolan CV, Borsboom D. Interpretational issues with the bifactor model: a commentary on “Defining the p-Factor: An Empirical Test of Five Leading Theories” by Southward, Cheavens, and Coccaro. Psychol Med. 2023 May;53(7):2744–7.

50. Chatzinakos C, Pernia CD, Morrison FG, Iatrou A, McCullough KM, Schuler H, et al. Single-Nucleus Transcriptome Profiling of Dorsolateral Prefrontal Cortex: Mechanistic Roles for Neuronal Gene Expression, Including the 17q21.31 Locus, in PTSD Stress Response. Am J Psychiatry. 2023 Oct 1;180(10):739–54.

51. Bountress KE, Bustamante D, Subbie-Saenz De Viteri S, Chatzinakos C, Sheerin C, Daskalakis NP, et al. Differences in genetic correlations between posttraumatic stress disorder and alcohol-related problems phenotypes compared to alcohol consumption-related phenotypes. Psychol Med. 2023 Sep;53(12):5767–77.

52. Whitfield JB, Zhu G, Madden PA, Neale MC, Heath AC, Martin NG. The Genetics of Alcohol Intake and of Alcohol Dependence. Alcohol Clin Exp Res. 2004;28(8):1153–60.

53. Barr PB, Bigdeli TB, Meyers JL, Peterson RE, Sanchez-Roige S, Mallard TT, et al. Correlates of Risk for Disinhibited Behaviors in the Million Veteran Program Cohort. MedRxiv Prepr Serv Health Sci. 2023 Jul 11;2023.03.22.23286865.

54. Savage JE, Barr PB, Phung T, Lee YH, Zhang Y, McCutcheon VV, et al. Genetic Heterogeneity Across Dimensions of Alcohol Use Behaviors. Am J Psychiatry. 2024 Nov 1;181(11):1006–17.

55. Hatoum AS, Colbert SMC, Johnson EC, Huggett SB, Deak JD, Pathak G, et al. Multivariate genome-wide association meta-analysis of over 1 million subjects identifies loci underlying multiple substance use disorders. Nat Ment Health. 2023 Mar;1(3):210–23.

56. Sidorenko J, Couvy-Duchesne B, Kemper KE, Moen GH, Bhatta L, Åsvold BO, et al. Genetic architecture reconciles linkage and association studies of complex traits. Nat Genet. 2024 Nov;56(11):2352–60.

57. Karlsson Linnér R, Biroli P, Kong E, Meddens SFW, Wedow R, Fontana MA, et al. Genome-wide association analyses of risk tolerance and risky behaviors in over 1 million individuals identify hundreds of loci and shared genetic influences. Nat Genet. 2019 Feb;51(2):245–57.

58. Brazel DM, Jiang Y, Hughey JM, Turcot V, Zhan X, Gong J, et al. Exome Chip Meta-analysis Fine Maps Causal Variants and Elucidates the Genetic Architecture of Rare Coding Variants in Smoking and Alcohol Use. Biol Psychiatry. 2019 Jun 1;85(11):946–55.

59. Rajabi A, Khosravi P, Motevalian SA, Farjam M, Shojaei A. The association between polymorphism of the BDNF gene and cigarette smoking in the Iranian population. J Gene Med. 2018 Oct;20(10–11):e3052.

60. Yang S, Wang F, Sun L, Liu X, Li S, Chen Y, et al. The effects of BDNF rs6265 and FGF21 rs11665896 polymorphisms on alcohol use disorder-related impulsivity in Han Chinese adults. Front Psychiatry. 2024;15:1339558.

61. Honarmand H, Bonyadi M, Rafat S, Mahdavi R, Aliasghari F. Association study of the BDNF gene polymorphism (G196A) with overweight/obesity among women from Northwest of Iran. Egypt J Med Hum Genet. 2021 Feb 1;22:7.

62. He L, Loika Y, Park Y, Bennett DA, Kellis M, Kulminski AM. Exome-wide age-of-onset analysis reveals exonic variants in ERN1 and SPPL2C associated with Alzheimer’s disease. Transl Psychiatry. 2021 Feb 26;11(1):1–18.

63. Colbert SMC, Funkhouser SA, Johnson EC, Morrison CL, Hoeffer CA, Friedman NP, et al. Novel characterization of the multivariate genetic architecture of internalizing psychopathology and alcohol use. Am J Med Genet B Neuropsychiatr Genet. 2021;186(6):353–66.

64. Stephenson M, Lannoy S, Edwards AC. Shared genetic liability for alcohol consumption, alcohol problems, and suicide attempt: Evaluating the role of impulsivity. Transl Psychiatry. 2023 Mar 10;13(1):1–8.

65. Carmiol N, Peralta JM, Almasy L, Contreras J, Pacheco A, Escamilla MA, et al. Shared genetic factors influence risk for bipolar disorder and alcohol use disorders. Eur Psychiatry. 2014 Jun;29(5):282–7.

66. Gasperi M, Panizzon M, Goldberg J, Buchwald D, Afari N. Posttraumatic Stress Disorder and Chronic Pain Conditions in Men: A Twin Study. Biopsychosoc Sci Med. 2021 Mar;83(2):109.

67. Yeung EW, Craggs JG, Gizer IR. Comorbidity of Alcohol Use Disorder and Chronic Pain: Genetic Influences on Brain Reward and Stress Systems. Alcohol Clin Exp Res. 2017 Nov;41(11):1831–48.

68. Dick DM, Balcke E, McCutcheon V, Francis M, Kuo S, Salvatore J, et al. The collaborative study on the genetics of alcoholism: Sample and clinical data. Genes Brain Behav. 2023 Oct;22(5):e12860.

