## Supplemental info for "Multivariate genome-wide association study (GWAS) of PTSD, Alcohol Use and Alcohol Use Disorders"

Supplemental Methods

Our bioinformatic pipeline had numerous steps. First, we tested gene-level and gene-set enrichment analysis using MAGMA (version 1.08, (35)), and its recent intersessions (FUMA version 1.3.6, (36)). In all the MAGMA-based analyses, SNPs were annotated to the 20,260 coding genes from Ensembl v92, with a 1 kb window for both sides (i.e., start and end). We corrected all MAGMA-based associations for multiple-testing using a Bonferroni correction.

The JEPEGMIX2-P software (37), using default parameters, was employed to conduct a Transcriptome-Wide Association Study (TWAS), utilizing 13 brain-specific genetically regulated expression (GReX) models from GTEx v8. This analysis leveraged genome-wide association study (GWAS) summary statistics to estimate gene-trait associations (GTAs) based on predicted GReX. A within-tissue Bonferroni correction was applied to identify statistically significant TWAS genes. For this study, the within tissue FDR significance was applied to detect significant TWAS genes.

Gene set enrichment analysis (GSEA) (38,39) implementation in R (FGSEA) was used to test concordance of differential expression analysis results with ~32K gene expression signatures (i.e., gene sets) from Molecular Signatures Database (MSigDB, datasets: http://software.broadinstitute.org/gsea/msigdb). For each input dataset, all genes were used for GSEA, and they were ranked based on Sign (Zscore) * -log10 (P-Value). The gene set size range was set between 15 and 501 genes. The GSEA returns, for each pathway, the enrichment P-value and the corresponding FDR adjusted P-value based on a permutation test (i.e., control for the number of the different gene sets tested for enrichment.), the Enrichment score (ES), the Normalized enrichment score (NES; i.e., enrichment score normalized to mean enrichment of random samples of the same pathway size), the nMoreExtreme (i.e., the number of times a random pathway had a larger ES), the size (i.e., size of the pathways after removing the genes not present) and the leadingEdge (i.e., the leading edge genes for each pathway that drive the enrichment). The number of pathways with FDR-adjusted p-value < 0.05 were reported for all pathway results. The ~32K gene expression signatures are organized into nine broader pathway categories, offering a structured approach to understanding the relationship between gene genes implicated by GWAS and canonical biological pathways. A detailed description of the pathways and all of the post-GWAS analyses can be found in the Supplementary Information.

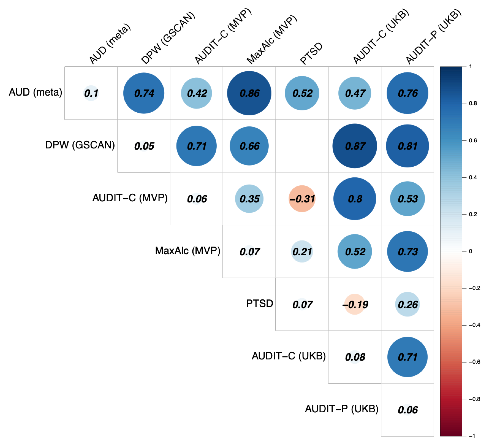

Supplemental Figure 1: Genetic Correlations Between GenomicSEM Indicators

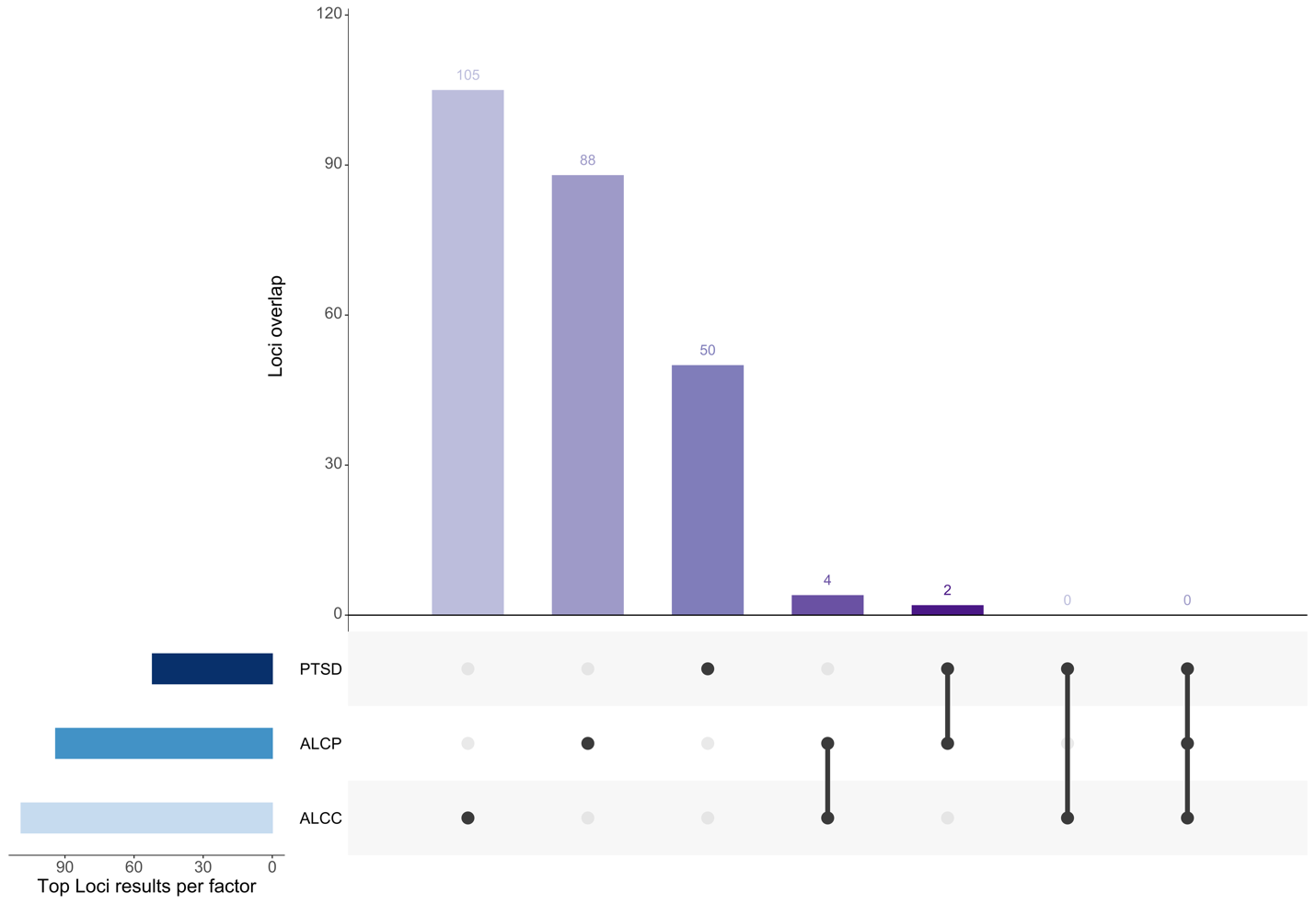

Supplemental Figure 2: Overlap in Top Loci from Multivariate GWAS

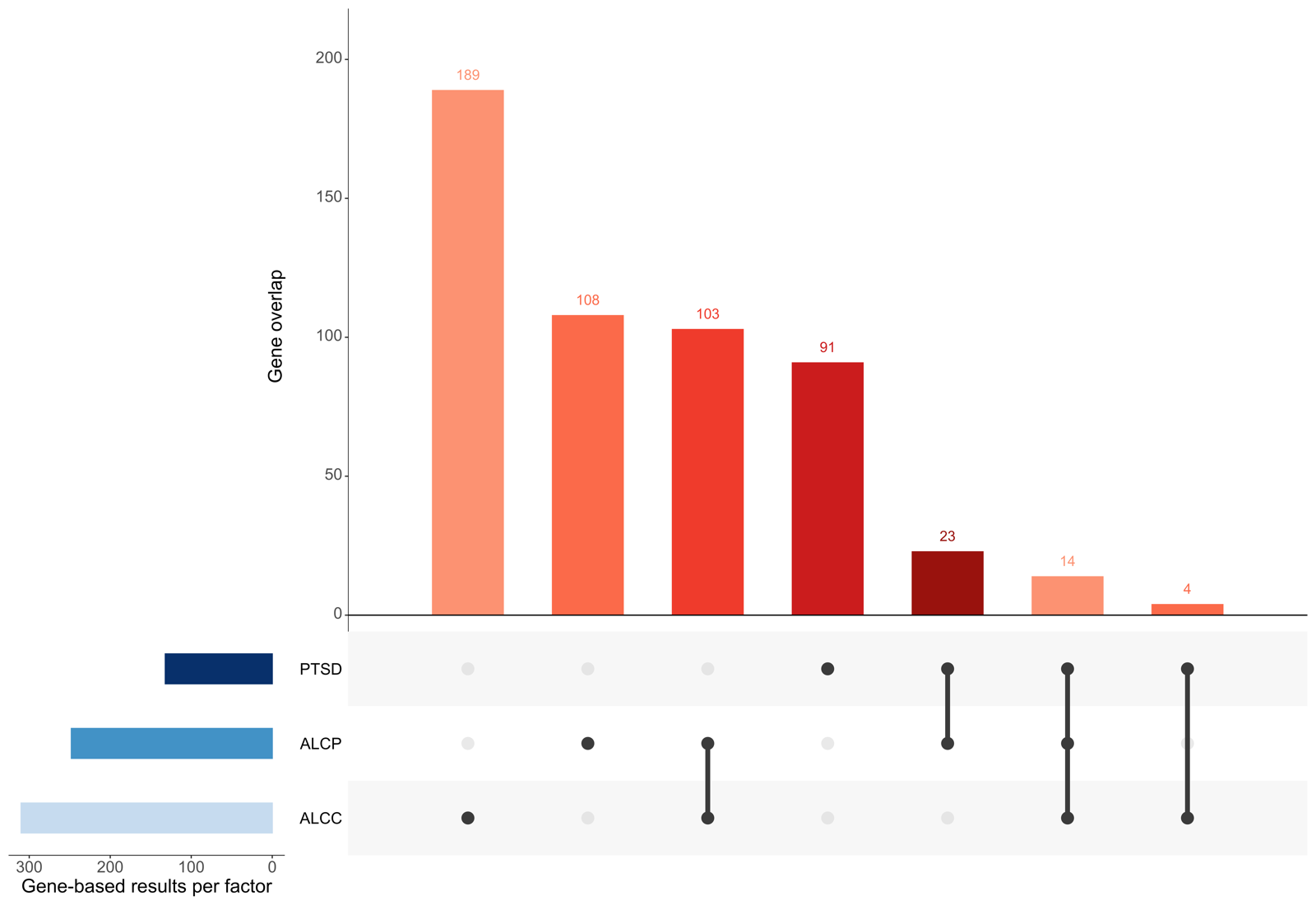

Supplemental Figure 3: Overlap in Gene-based Results from Multivariate GWAS

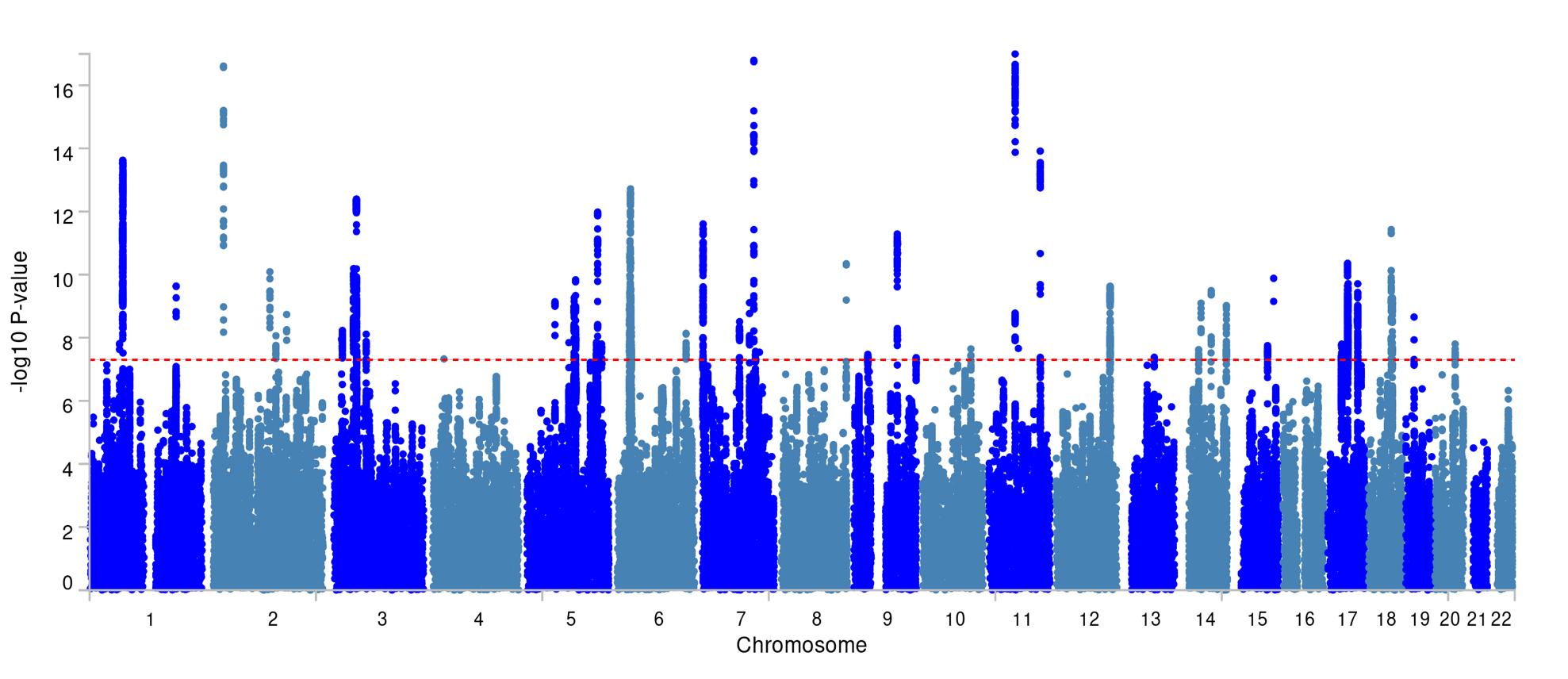

Supplemental Figure 4: Manhattan Plot for PTSD-factor GWAS

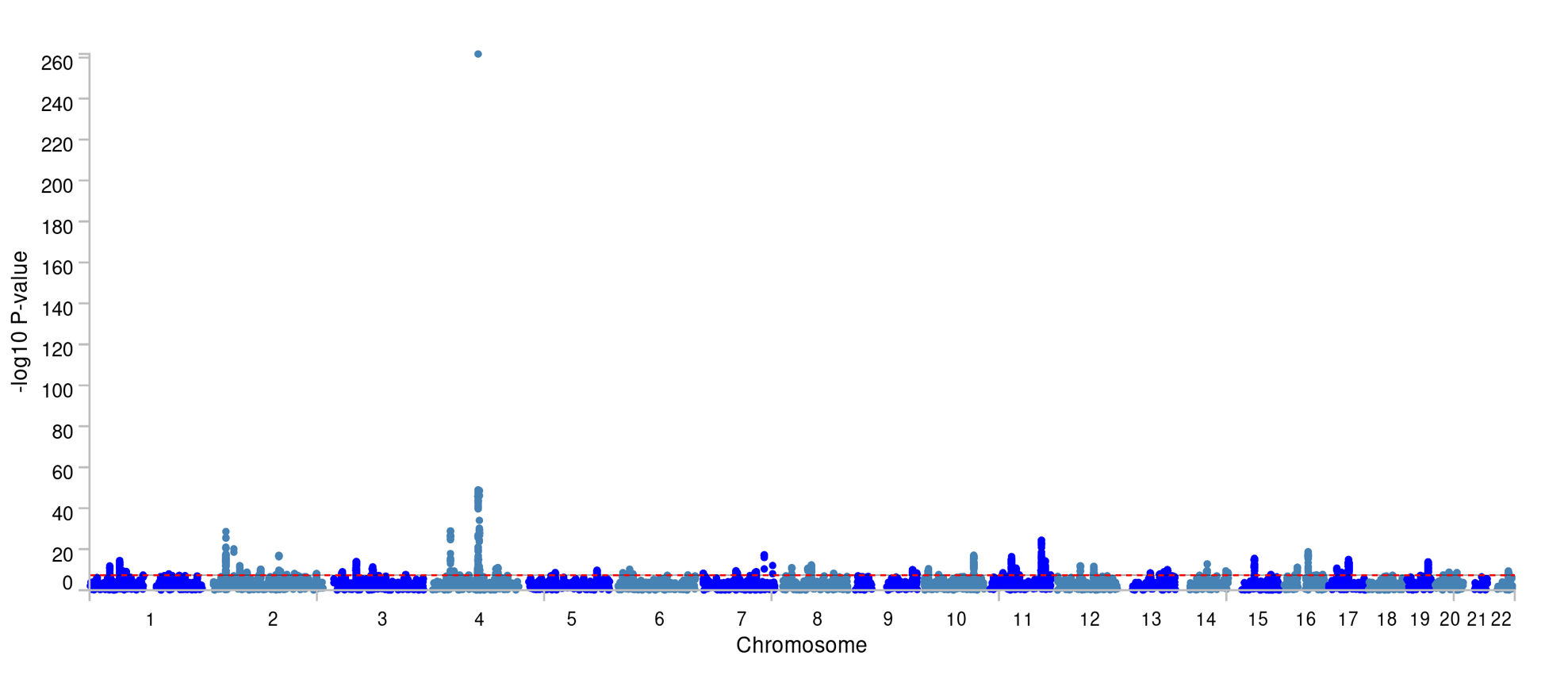

Supplemental Figure 5: Manhattan Plot for ALCP-factor GWAS

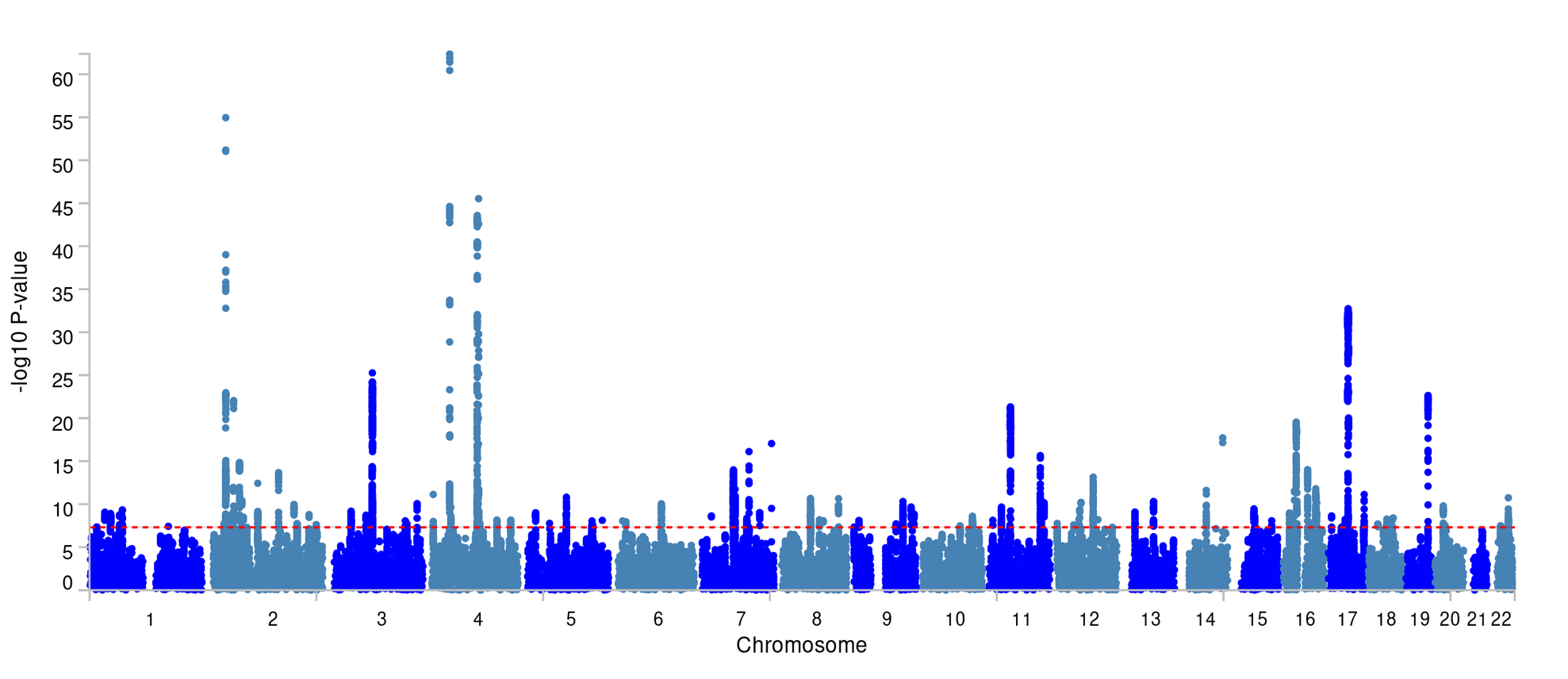

Supplemental Figure 6: Manhattan Plot for ALCC-factor GWAS

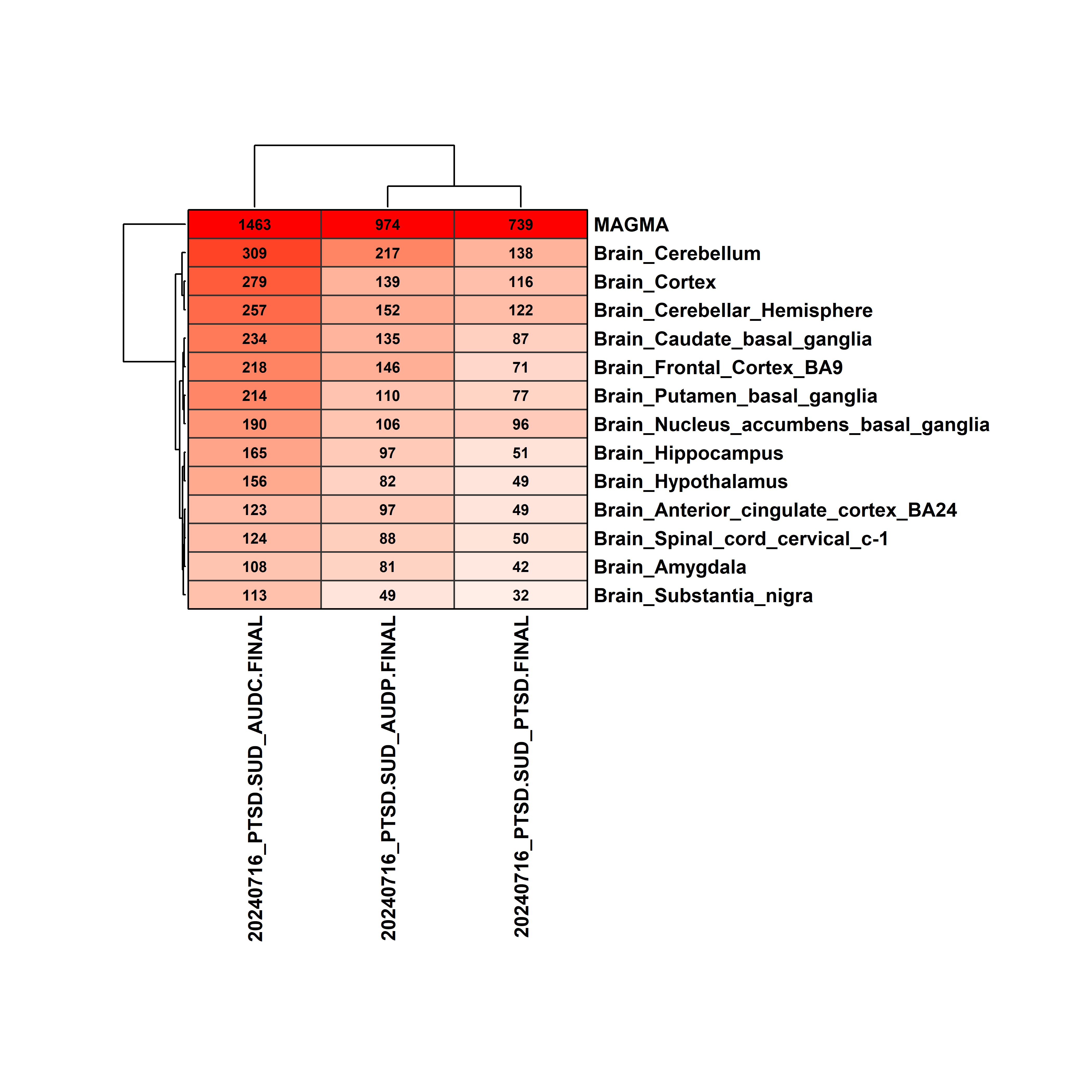

Supplemental Figure 7: TWAS Results for GenomicSEM Factors

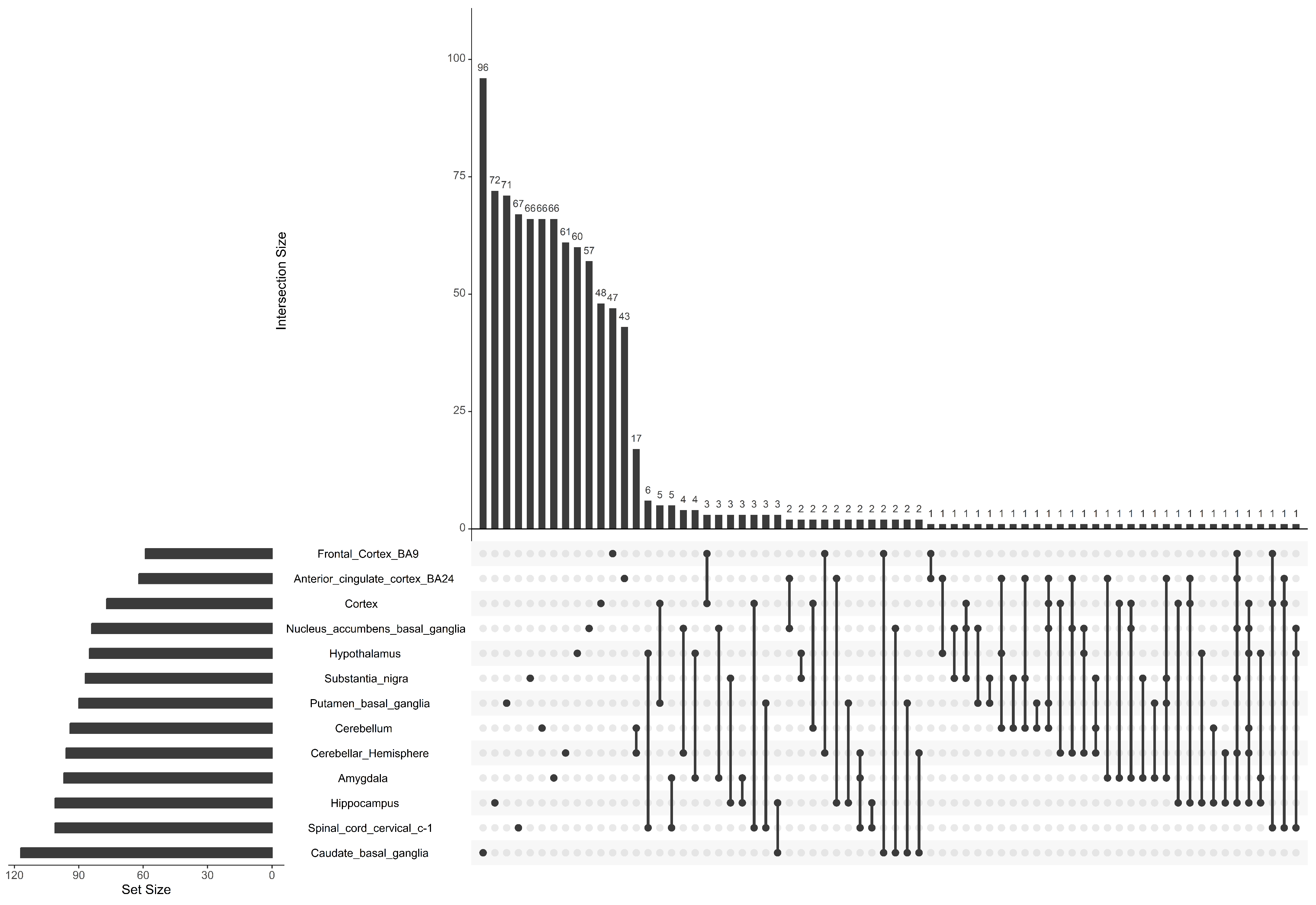

Supplemental Figure 8: Significant Pathway (FDR<0.05) overlap for PTSD-factor GWAS across the 13 GTeX-Brain Tissues

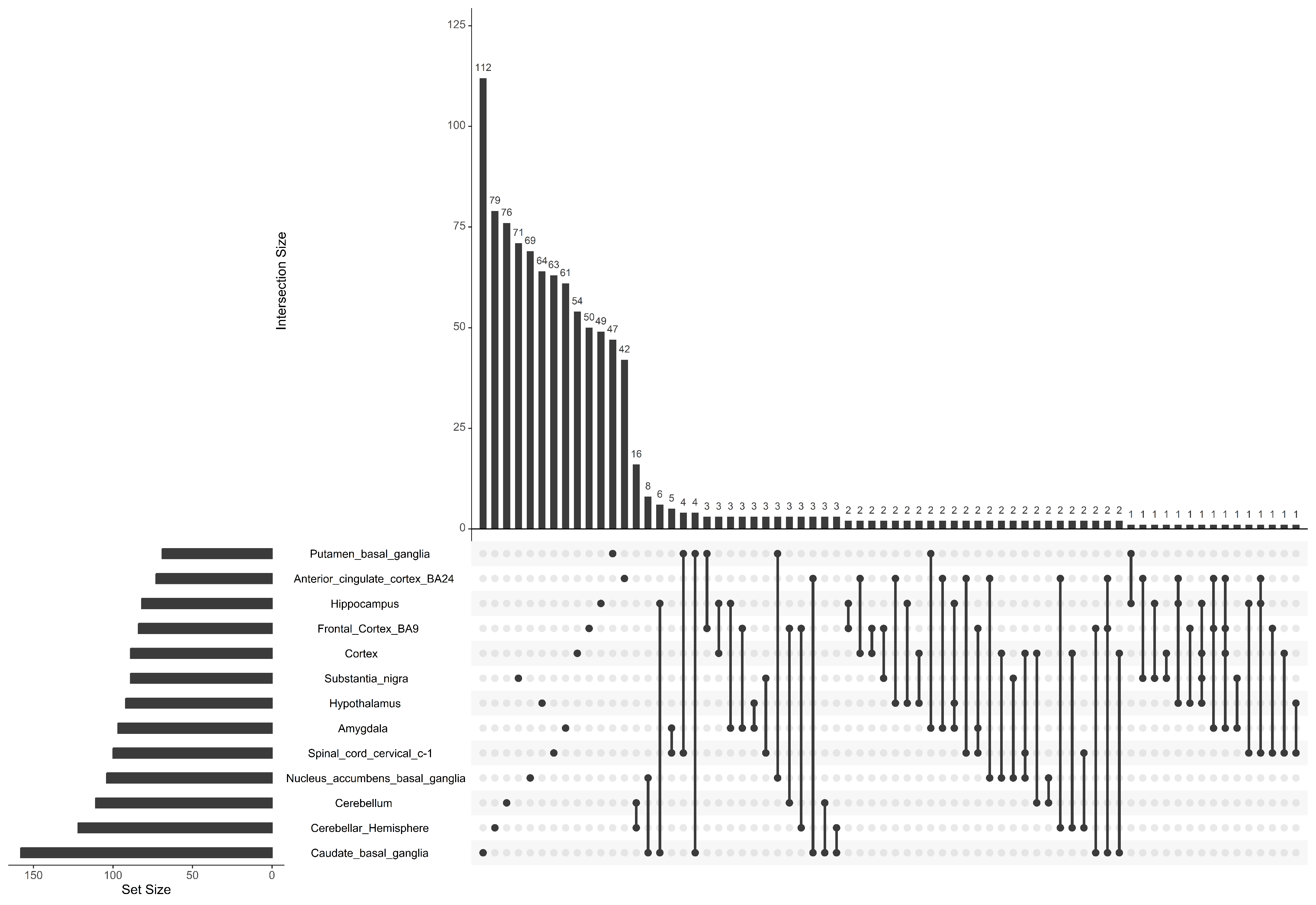

Supplemental Figure 9: Significant Pathway (FDR<0.05) overlap for original PTSD GWAS across the 13 GTeX-Brain Tissues

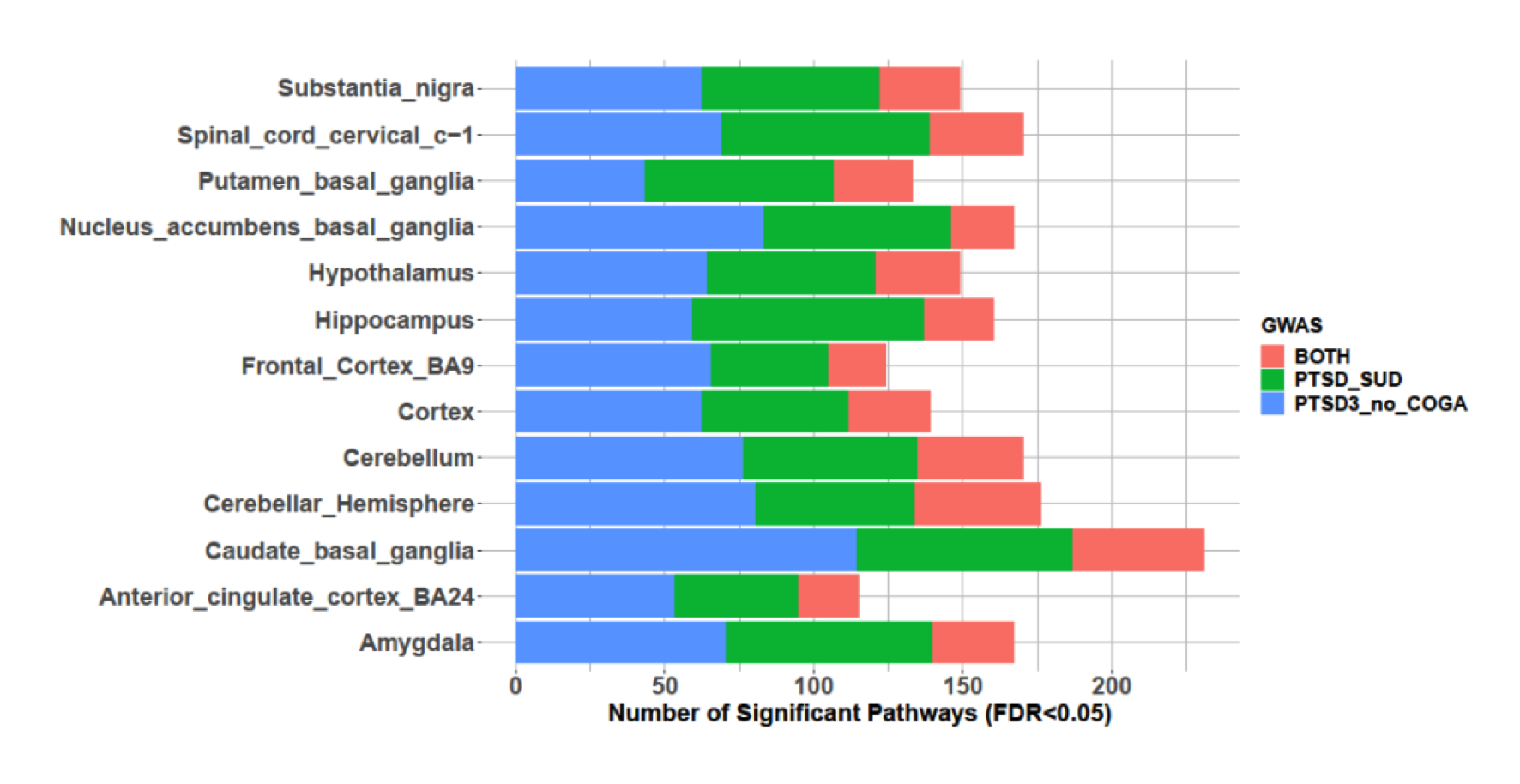

Supplemental Figure 10: Significant Pathway (FDR<0.05) overlap for PTSD-factor GWAS and original PTSD GWAS across the 13 GTeX-Brain Tissues

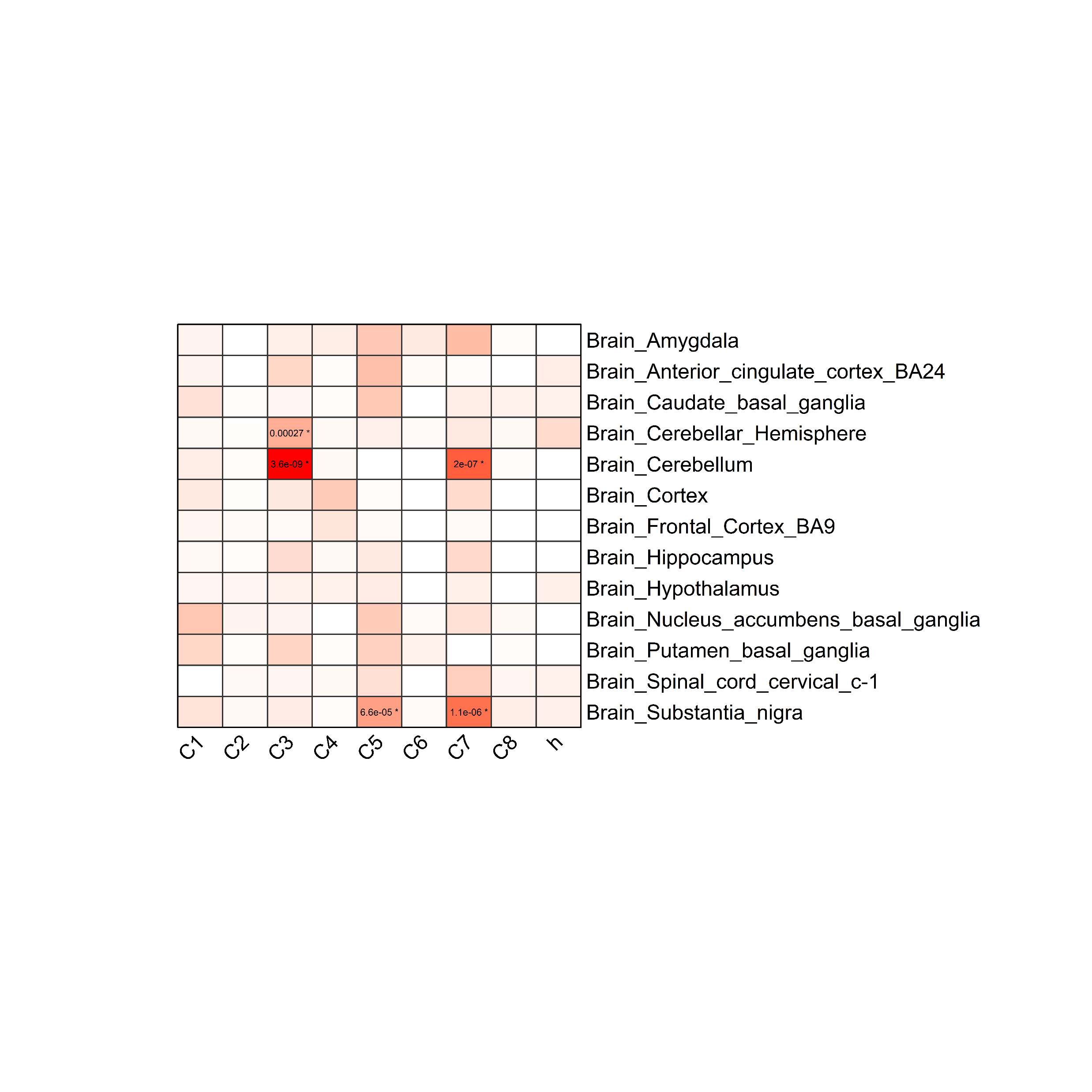

Supplemental Figure 11: Phisher exact test for the Significant Pathway (FDR<0.05) of PTSD-factor GWAS across the 9 categories

1. Hallmark Gene Sets (H): These are coherently expressed gene signatures that aggregate multiple gene sets representing well-defined biological processes and states. They are derived from a large number of gene sets to provide a focused insight into key biological mechanisms.

2. Positional Gene Sets (C1): These correspond to genes located in specific human chromosome cytogenetic bands, focusing on genomic regions that may have a role in diseases or traits, such as those implicated in PTSD-SUD.

3. Curated Gene Sets (C2): These gene sets are collected from expert-validated databases, publications, and pathway knowledge sources. They represent known biological pathways and molecular functions relevant to PTSD-SUD.

4. Regulatory Target Gene Sets (C3): These sets are based on predicted transcription factor binding sites and microRNA targets, showing how gene expression may be regulated at a transcriptional or post-transcriptional level, potentially affecting PTSD-SUD risk.

5. Computational Gene Sets (C4): Defined by large cancer-oriented expression datasets, these sets are mined for patterns in gene expression that can be applied across different diseases, providing a broader context of gene activity.

6. Ontology Gene Sets (C5): These consist of genes grouped under the same ontology term, such as biological processes, molecular functions, or cellular components. This category is useful for determining the functional roles of genes in PTSD-SUD.

7. Oncogenic Signature Gene Sets (C6): Defined from cancer perturbation data, these gene sets highlight pathways influenced by oncogenic processes and provide insight into broader disease mechanisms, including PTSD-SUD.

8. Immunologic Signature Gene Sets (C7): These gene sets represent immune-related states and cell perturbations. Given the involvement of the immune system in stress and trauma responses, this category is particularly relevant to PTSD-SUD.

9. Cell Type Signature Gene Sets (C8): Curated from single-cell sequencing data, these gene sets identify specific cell types, offering insights into how distinct cell populations may contribute to PTSD-SUD pathology.

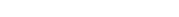

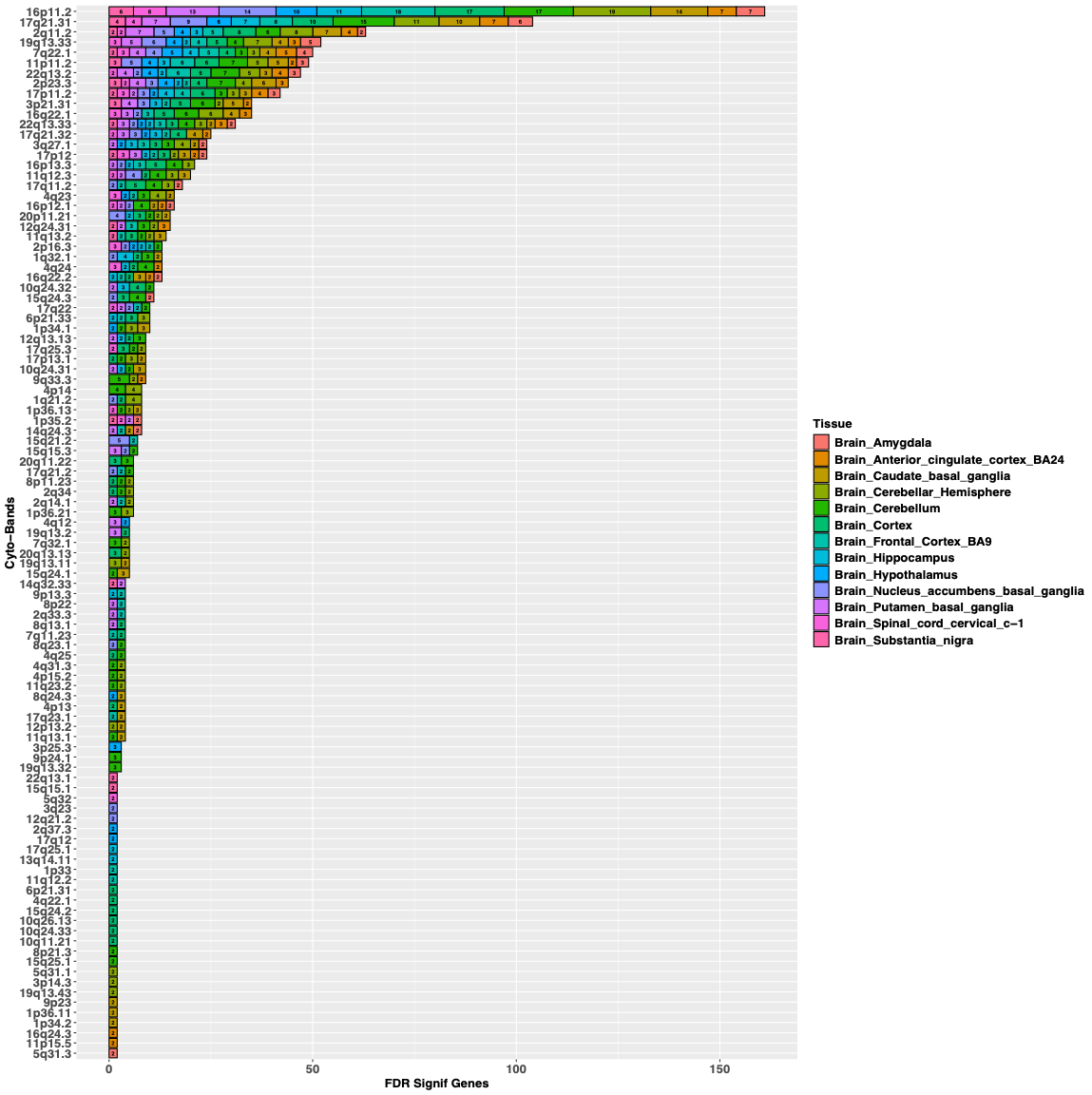

Supplemental Figure 12: FDR Significant Genes by Band for ALCC

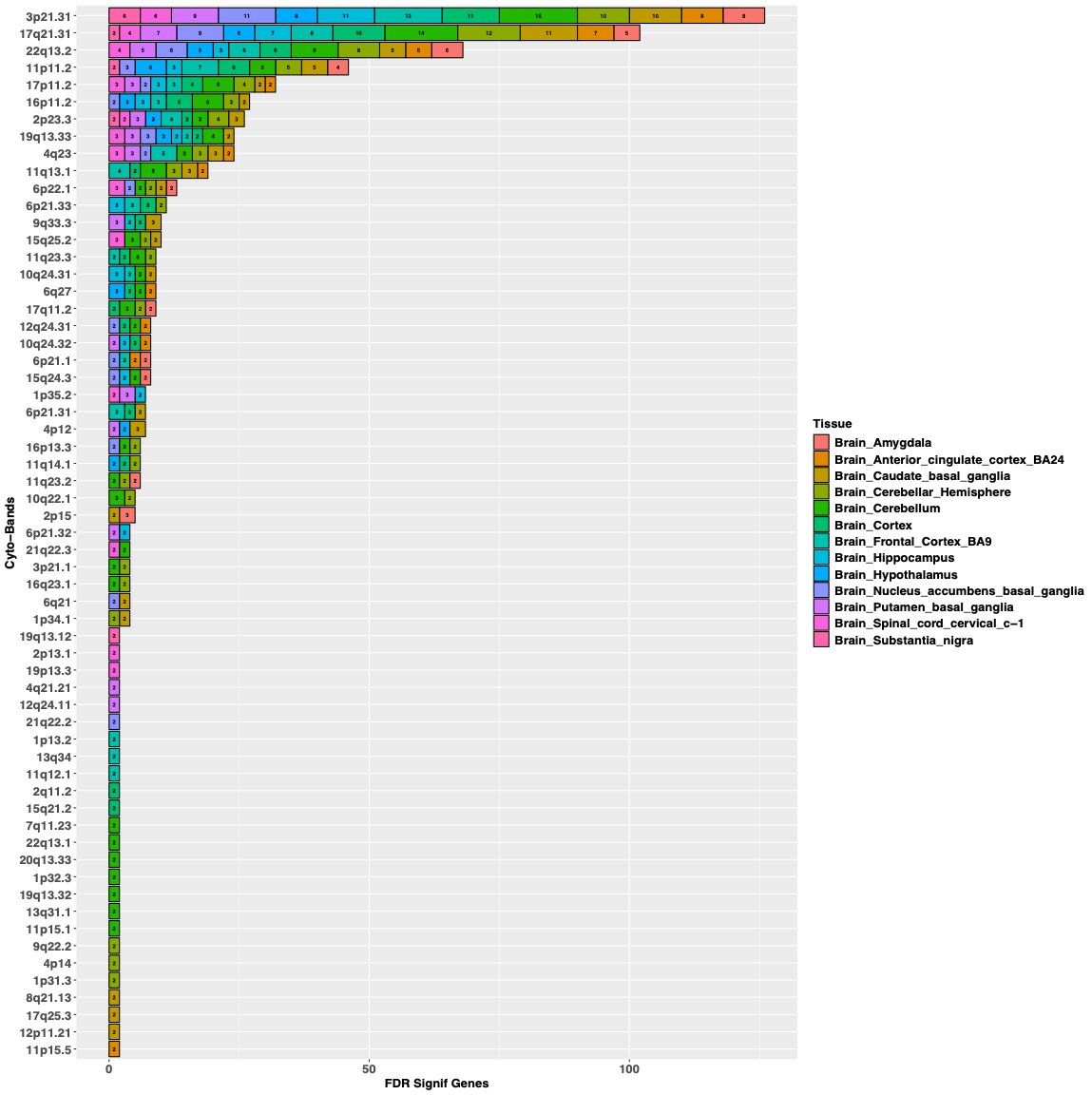

Supplemental Figure 13: FDR Significant Genes by Band for ALCP

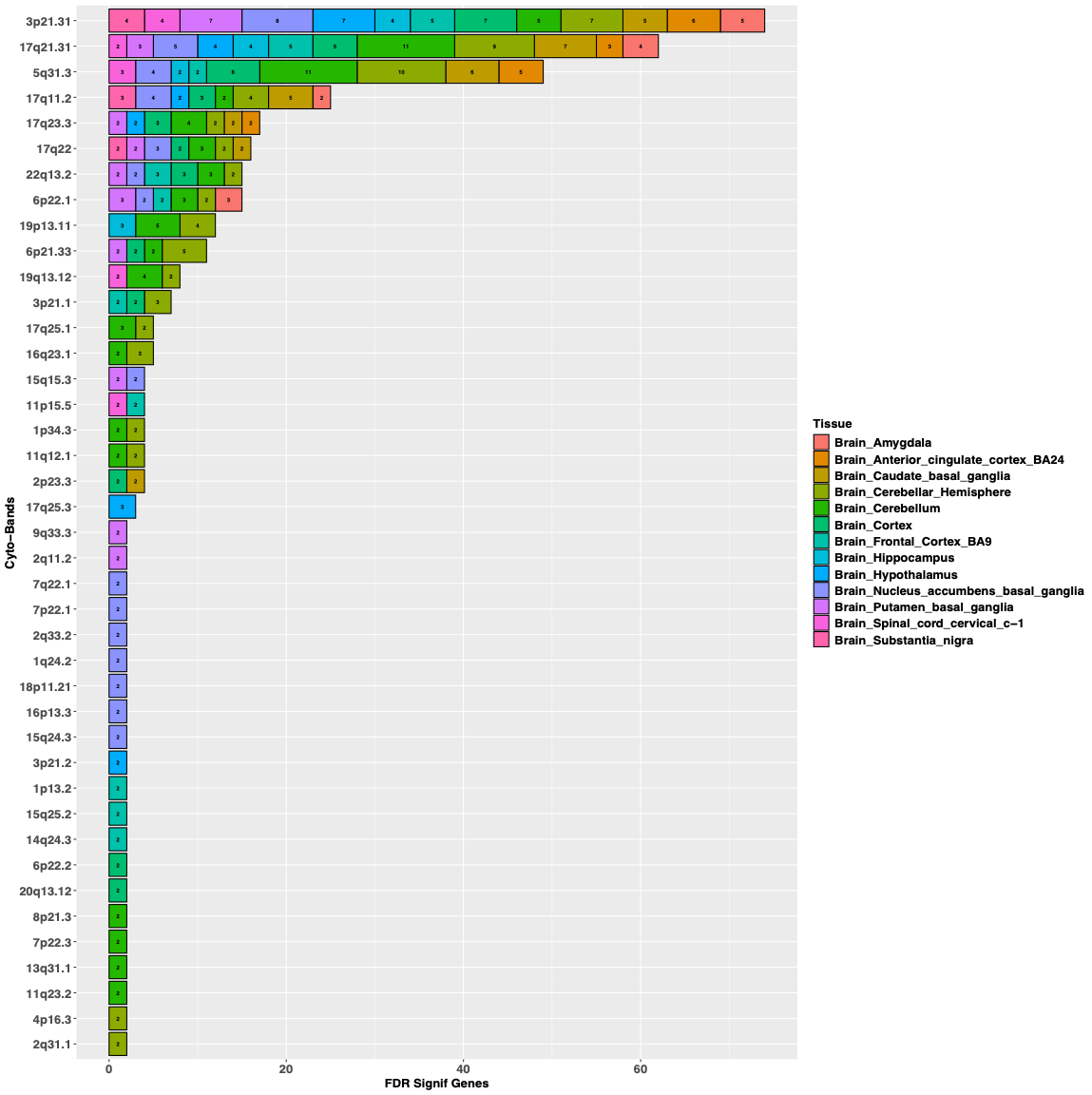

Supplemental Figure 14: FDR Significant Genes by Band for PTSD
